# Actionable druggable genome-wide Mendelian randomization identifies repurposing opportunities for COVID-19

**DOI:** 10.1101/2020.11.19.20234120

**Authors:** Liam Gaziano, Claudia Giambartolomei, Alexandre C Pereira, Anna Gaulton, Daniel C Posner, Sonja A Swanson, Yuk-Lam Ho, Sudha K Iyengar, Nicole M Kosik, Marijana Vujkovic, David R Gagnon, A Patrícia Bento, Pedro Beltrao, Inigo Barrio-Hernandez, Lars Rönnblom, Niklas Hagberg, Christian Lundtoft, Claudia Langenberg, Maik Pietzner, Dennis Valentine, Elias Allara, Praveen Surendran, Stephen Burgess, Jing Hua Zhao, James E Peters, Bram P Prins, John Danesh, Poornima Devineni, Yunling Shi, Kristine E Lynch, Scott L DuVall, Helene Garcon, Lauren O Thomann, Jin J Zhou, Bryan R Gorman, Jennifer E Huffman, Christopher J O’Donnell, Philip S Tsao, Jean C Beckham, Saiju Pyarajan, Sumitra Muralidhar, Grant D Huang, Rachel Ramoni, Adriana M Hung, Kyong-Mi Chang, Yan V Sun, Jacob Joseph, Andrew R Leach, Todd L Edwards, Kelly Cho, J Michael Gaziano, Adam S Butterworth, Juan P Casas, on behalf VA Million Veteran Program COVID-19 Science Initiative

**Affiliations:** Massachusetts Veterans Epidemiology Research and Information Center (MAVERIC), VA Boston Healthcare System, Boston, MA, USA; BHF Cardiovascular Epidemiology Unit, Department of Public Health and Primary Care, University of Cambridge, Cambridge, UK; Central RNA Lab, Istituto Italiano di Tecnologia, Genova, Italy; Department of Pathology and Laboratory Medicine, David Geffen School of Medicine, University of California Los Angeles, Los Angeles, CA, USA; Laboratory of Genetics and Molecular Cardiology, Heart Institute, University of Sao Paulo, Sao Paulo, Brazil; Genetics Department, Harvard Medical School, Harvard University, Boston, MA, USA; Chemical Biology, European Molecular Biology Laboratory, European Bioinformatics Institute, Hinxton, UK; Department of Epidemiology, Erasmus Medical Center, Rotterdam, The Netherlands; Louis Stokes Cleveland VA Medical Center, Cleveland, OH, USA; Department of Population and Quantitative Health Sciences, Case Western Reserve University and Louis Stoke, Cleveland VA, Cleveland, OH, USA; The Corporal Michael J. Crescenz VA Medical Center, the University of Pennsylvania Perelman School of Medicine, Philadelphia, PA, USA; Medicine, Perelman School of Medicine, University of Pennsylvania, Philadelphia, PA, USA; Biostatistics, School of Public Health, Boston University, Boston, MA, USA; European Molecular Biology Laboratory, European Bioinformatics Institute, Hinxton, UK; Department of Medical Sciences, Uppsala University, Uppsala, Sweden; Berlin Insitute of Health, Charité University Medicine Berlin, Berlin, Germany; MRC Epidemiology Unit, Universityof Cambridge, Cambridge, UK; Institute of Health Informatics, University College London, London, UK; Health Data Research, University College London, London, UK; British Heart Foundation Centre of Research Excellence, University of Cambridge, Cambridge, UK; Health Data Research UK Cambridge, Wellcome Genome Campus and University of Cambridge, Cambridge, UK; Rutherford Fund Fellow, Department of Public Health and Primary Care, University of Cambridge, Cambridge, UK; MRC Biostatistics Unit, University of Cambridge, Cambridge, UK; Centre for Inflammatory Disease, Dept of Immunology and Inflammation, Imperial College, London, UK; Health Data Research UK, UK; VA Informatics and Computing Infrastructure, VA Salt Lake City Health Care System, Salt Lake City, UT, USA; Department of Internal Medicine, Epidemiology, University of Utah, Salt Lake City, UT, USA; Department of Epidemiology and Biostatistics, University of Arizona, Tucson, AZ, USA; Phoenix VA Health Care System, Phoenix, AZ, USA; Center for Population Genomics, Massachusetts Veterans Epidemiology Research and Information Center (MAVERIC), VA Boston Healthcare System, Boston, MA, USA; Cardiology, VA Boston Healthcare System, Boston, MA, USA; Medicine, Brigham and Women’s Hospital, Harvard Medical School, Boston, MA, USA; Epidemiology Research and Information Center (ERIC), VA Palo Alto Health Care System, Palo Alto, CA, USA; Department of Medicine, Stanford University School of Medicine, Palo Alto, CA, USA; MIRECC, Durham VA Medical Center, Durham, NC, USA; Department of Psychiatry and Behavioral Sciences, Duke University School of Medicine, Durham, NC, USA; Office of Research and Development, Department of Veterans Affairs, Washington, DC, USA; VA Tennessee Valley Healthcare System, Nashville, TN, USA; Nephrology & Hypertension, Vanderbilt University, Nashville, TN, USA; The Corporal Michael J. Crescenz VA Medical Center, Philadelphia, PA, USA; Department of Medicine, Perlman School of Medicine, University of Pennsylvania, Philadelphia, PA, USA; Atlanta VA Health Care System, Decatur, GA, USA; Department of Epidemiology, Emory University Rollins School of Public Health, Atlanta, GA, USA; Medicine, Cardiovascular, VA Boston Healthcare System and Brigham & Women’s Hospital, Boston, MA, USA; Department of Veterans Affairs, Tennessee Valley Healthcare System, Vanderbilt University, Nashville, TN, USA; Medicine, Epidemiology, Vanderbilt Genetics Institute, Vanderbilt University Medical Center, Nashville, TN, USA; Division of Aging, Brigham and Women’s Hospital, Harvard Medical School, Boston, MA, USA; National Institute for Health Research Blood and Transplant Research Unit in Donor Health and Genomics, University of Cambridge, Cambridge, UK; National Institute for Health Research Cambridge Biomedical Research Centre, University of Cambridge and Cambridge University Hospitals, Cambridge, UK

**Keywords:** Mendelian randomization, drug repurposing, COVID-19, actionable druggable genome

## Abstract

Drug repurposing provides a rapid approach to meet the urgent need for therapeutics to address COVID-19. To identify therapeutic targets relevant to COVID-19, we conducted Mendelian randomization (MR) analyses, deriving genetic instruments based on transcriptomic and proteomic data for 1,263 actionable proteins that are targeted by approved drugs or in clinical phase of drug development. Using summary statistics from the Host Genetics Initiative and the Million Veteran Program, we studied 7,554 patients hospitalized with COVID-19 and >1 million controls. We found significant Mendelian randomization results for three proteins (ACE2: *P=*1.6×10^−6^, IFNAR2: *P=*9.8×10^−11^, and IL-10RB: *P=*1.9×10^−14^) using *cis*-eQTL genetic instruments that also had strong evidence for colocalization with COVID-19 hospitalization. To disentangle the shared eQTL signal for *IL10RB* and *IFNAR2*, we conducted phenome-wide association scans and pathway enrichment analysis, which suggested that *IFNAR2* is more likely to play a role in COVID-19 hospitalization. Our findings prioritize trials of drugs targeting IFNAR2 and ACE2 for early management of COVID-19.

## INTRODUCTION

COVID-19 has caused a global pandemic resulting in excess mortality, stress on healthcare systems and economic hardship. Even if an efficacious vaccine against SARS-CoV-2 virus emerges in 2021, it could take several years to achieve herd immunity. Hence, there is a need to rapidly identify drugs that can minimize the burden of COVID-19. Although large randomized trials have begun to successfully identity drugs that can be repurposed to address COVID-19,^1,2^ most drugs evaluated so far have failed to show efficacy and have been largely confined to hospitalized or critically-ill patients. Therefore, it is pressing to identify additional drugs that can be repurposed for early management in COVID-19.

Large-scale human genetic studies are now widely used to inform drug development programs as drug target-disease pairs supported by human genetics have a greater odds of success in drug discovery pipelines.^3,4^ For example, identification of variants in *PCSK9* associated with lower risk of coronary disease led to the successful development of PCSK9 inhibitors, which are now licensed for prevention of cardiovascular events.^5^ The value of human genetics for drug discovery and development has also been realized for infectious diseases. Human genetic studies showed that genetic variation in the *CCR5* gene provides protection against infection by human immunodeficiency virus (HIV) type-1. These findings were key for the development of Maraviroc, an antagonist of CCR5, approved by the FDA for the treatment of patients with HIV-1.^6^

Genetic variants acting in “*cis*” on druggable protein levels or gene expression that encode druggable proteins can provide powerful tools for informing therapeutic targeting, as they mimic the on-target (beneficial or harmful) effects observed by pharmacological modification.^7^ Such Mendelian randomization (MR) analyses have been used to suggest repurposing opportunities for licensed drugs.^8,9^ MR analysis that focuses on actionable druggable genes, defined as genes that encode the protein targets of drugs that are licensed or in the clinical phase of drug development, could therefore serve as a fast and robust strategy to identify drug-repurposing opportunities to prevent the complications and mortality due to COVID-19.

To identify further potential repurposing opportunities to inform trials of COVID-19 patients, we conducted large-scale MR and colocalization analyses using gene expression and soluble protein data for 1,263 actionable druggable genes that encode protein targets for approved drugs or drugs in clinical development. By combining trans-ancestry genetic data from 7,554 hospitalized COVID-19 patients and more than 1 million population-based controls from the COVID-19 Host Genetics Initiative^10^ (HGI) and the Million Veteran Program^11^ (MVP), we provide support for two therapeutic strategies.

## RESULTS

### Overall analysis plan

**Figure 1** describes the overall scheme of the analyses. First, we identified all proteins that are therapeutic targets of approved or clinical-stage drugs. Next, we selected conditionally-independent genetic variants that act locally on plasma levels of these proteins or tissue-specific gene expression that encode these proteins. We proposed that these variants were instrumental variables and conduct two-sample MR analyses^12^ using a trans-ancestry meta-analysis of 7,554 cases from MVP and publicly available data (HGI outcome B2 from release 4 version 1, downloaded October 4^th^ 2020, **Supplementary Table 1**). Given that all MR analyses relies on several assumptions, some^13^ unverifiable, we conducted a multi-stage strategy to minimize confounding and biases. For MR results that passed our significance threshold after accounting for multiple testing, we performed colocalization to ensure MR results were not due to confounding by linkage disequilibrium (LD). Those with evidence of colocalization were investigated further using an independent proteomics platform (Olink). Finally, we conducted phenome-wide scans and pathway enrichment of relevant variants to reduce risks of horizontal pleiotropy and other biases due to MR violations as well as to understand potential biological mechanisms.

**Table 1.**
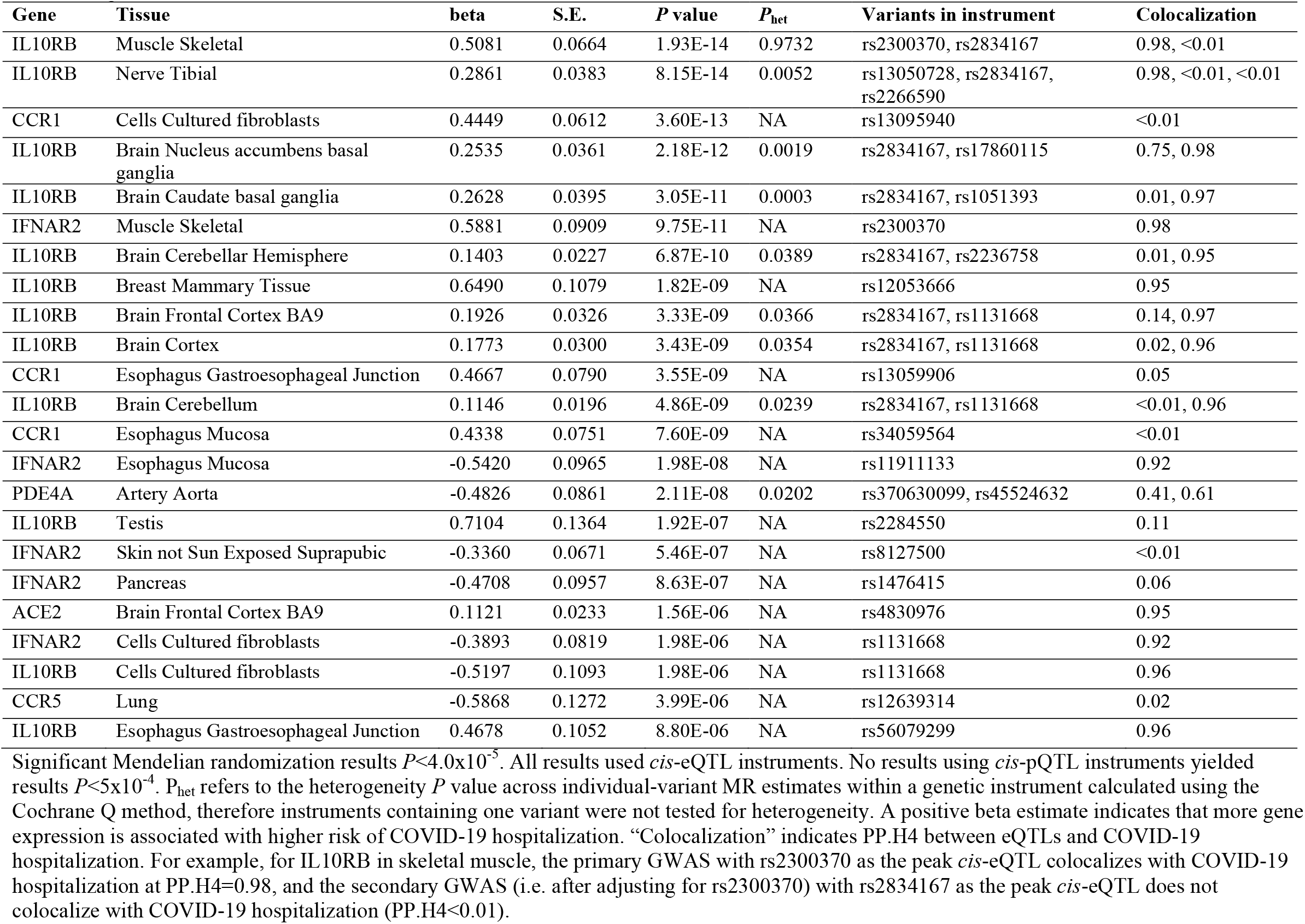
Significant (*P*<4.0×10^−5^) MR results.

**Figure 1.**
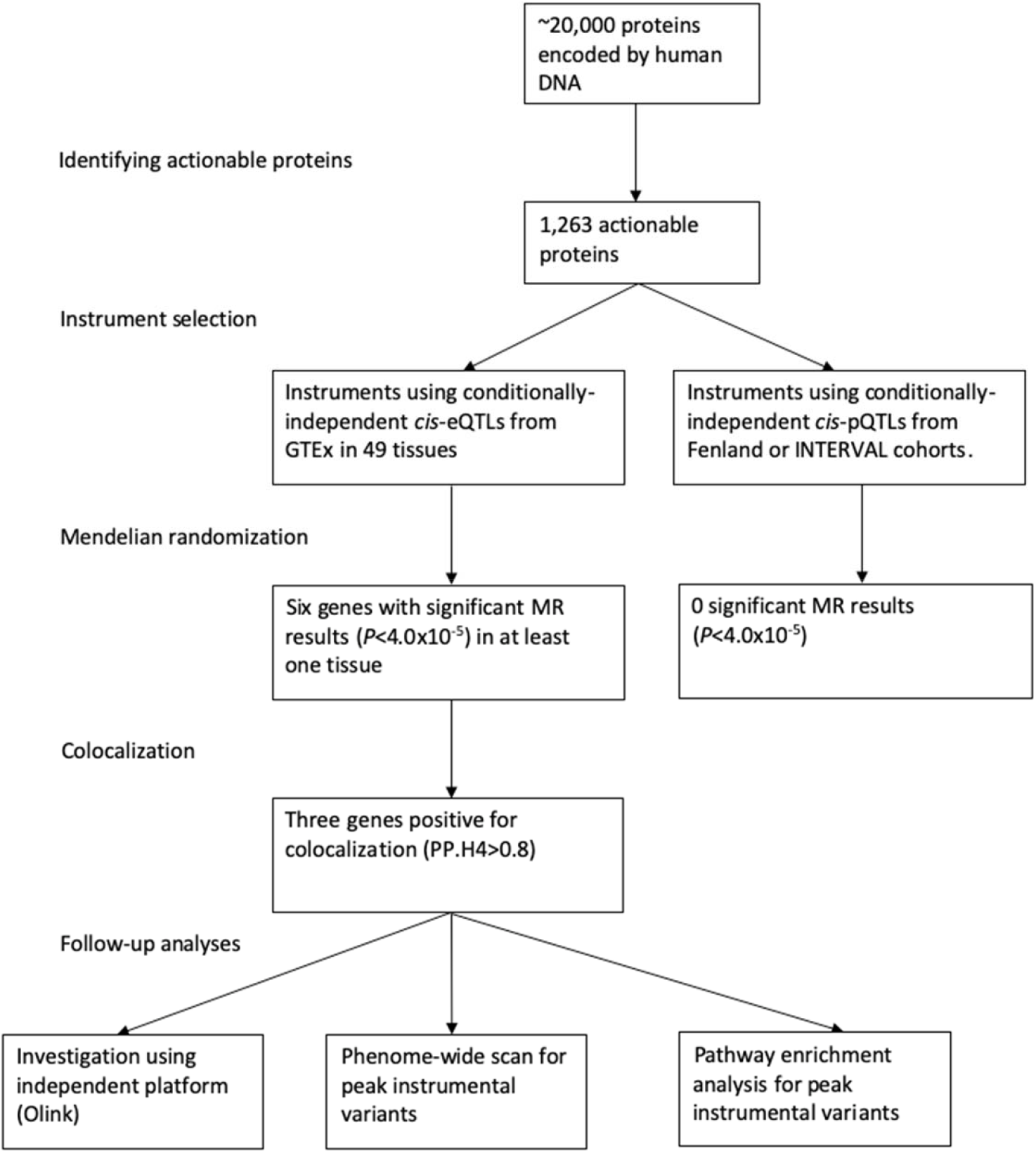
Outline of the analyses performed. Using multiple data sources, this study tested *cis*-pQTL and *cis*-eQTL proposed instruments for actionable druggable proteins against COVID-19 hospitalization summary statistics meta-analyzed from the Host Genetics Initiative and the Million Veteran program. Significant MR associations that also showed evidence for colocalization were investigated further with an independent platform (Olink), phenome-wide scans of relevant variants, and pathway enrichment.

### Actionable druggable proteins

Using data available in ChEMBL version 26, we identified 1,263 human proteins as ‘actionable’ (i.e. therapeutic targets of approved or clinical-stage drugs) (**Supplementary Table 2**). Of these, we noted 700 proteins that are targets for drugs with potential relevance to COVID-19 from cell-based screening, registers of clinical trials against COVID-19 or approved immunomodulatory/anticoagulant drugs (given the clear role of these pathways in COVID-19 outcomes), or have biological evidence for the role of the protein in SARS-CoV-2 infection (**Supplementary Table 3**).

### Genetic proposed instruments for actionable druggable proteins

Using GTEx version 8 (V8)^14^, we identified all conditionally-independent expression quantitative trait loci (eQTLs) in 49 tissues that act in *cis* (within 1 Mb on either side of the encoded gene), that covered 1,016 of the 1,263 druggable genes in at least one tissue (**Supplementary Table 2 and 4**). We also selected *cis*-pQTLs for plasma proteins measured using the SomaScan platform in 3,301 participants of the INTERVAL study^15^ (**Supplementary Table 5**) and 10,708 Fenland cohort participants^16^ (**Supplementary Table 6**) that covered a total of 67 proteins. In total 1,021 proteins had genetic proposed instruments using either eQTLs or pQTLs, and 62 had proposed instruments using both.

### Mendelian randomization and colocalization

Using our (eQTL and pQTL) proposed instruments, we performed two-sample MR on trans-ancestry summary statistics for hospitalized COVID-19 cases from MVP and HGI (**Supplementary Table 1)**. Using GTEx *cis*-eQTLs as proposed instruments, we found significant (*P*<4.0×10^−5^, 0.05 Bonferroni-corrected for 1,263 proteins) MR results for six genes (*IL10RB, CCR1, IFNAR2, PDE4A, ACE2* and *CCR5)* in at least one tissue (**Table 1)**, and four additional genes (*CA5B, CA9, NSTN* and *SLC9A3*) with suggestive MR results (*P*<5×10^−4^ and *P*>4×10^−5^, **Figure 2)**. No proposed instruments involving *cis*-pQTLs reached our suggestive threshold in any of the analyses. For three significant genes (*IL10RB, IFNAR2, ACE2*) there was strong evidence of colocalization (posterior probability of shared causal variant across two traits - hypothesis 4 [PP.H4] >0.8) between at least one proposed instrumental variant and our trans-ancestry meta-analysis of COVID-19 hospitalization (**Table 1**). Beta-coefficients of MR estimates for *ACE2* were positive in all tissues (**Table 1**), meaning higher *ACE2* expression is associated with higher risk of COVID-19 hospitalization. MR beta-coefficients for *IFNAR2* and *IL10RB* were negative and positive, respectively, in all tissues except one for each gene (skeletal muscle for *IFNAR2*; cultured fibroblasts for *IL10RB*; **Table 1**).

**Figure 2.**
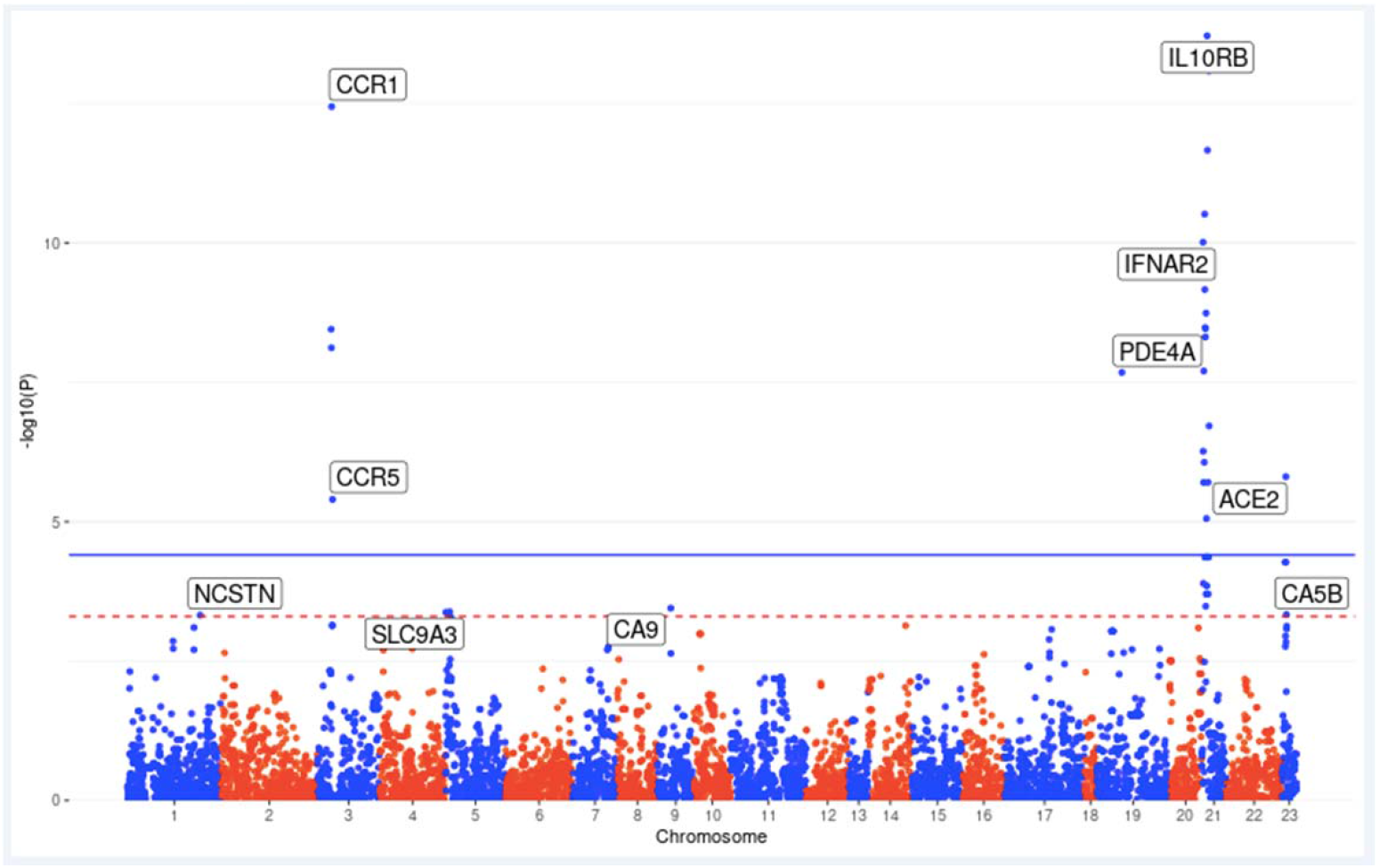
Manhattan plot of results from actionable druggable genome-wide Mendelian randomization analysis. Blue solid line indicates the *P* value threshold for significance (*P* <4.0×10^−5^, 0.05 Bonferroni-corrected for 1,263 actionable druggable genes) and red dashed line indicates the suggestive (*P*<5×10^−4^) threshold. Genes are labeled by their most significant MR association. For example, the results for *IL10RB* is most significant with *cis*-eQTL proposed instruments derived in skeletal muscle tissue, which is the point labeled. Results are plotted by the gene start position. All MR results with *P* value less than 5×10^−4^ used the GTEx *cis*-eQTLs as proposed instruments.

### IL10RB and IFNAR2

Interferon alpha receptor 2 (IFNAR2) and interleukin 10 receptor beta (IL-10RB) both act as receptors for interferons (IFN). IFNAR2 forms a complex with IFNAR1, which together act as a receptor for type I IFN (IFN-α, β, ω, κ, ε), while IL-10RB acts as a receptor for type III IFN (IFN-λ) when complexed with interferon lambda receptor-1 (IFNLR1)^17^, or IL-10 when complexed with IL-10RA. IL-10RB and IFNAR2 are encoded by adjacent genes and some *cis*-eQTLs for *IL10RB* are also *cis*-eQTLs for *IFNAR2* (**Supplementary Table 7, Figure 3**), making it difficult to determine which gene may be responsible for the association with COVID-19 and requiring further investigation.

**Figure 3.**
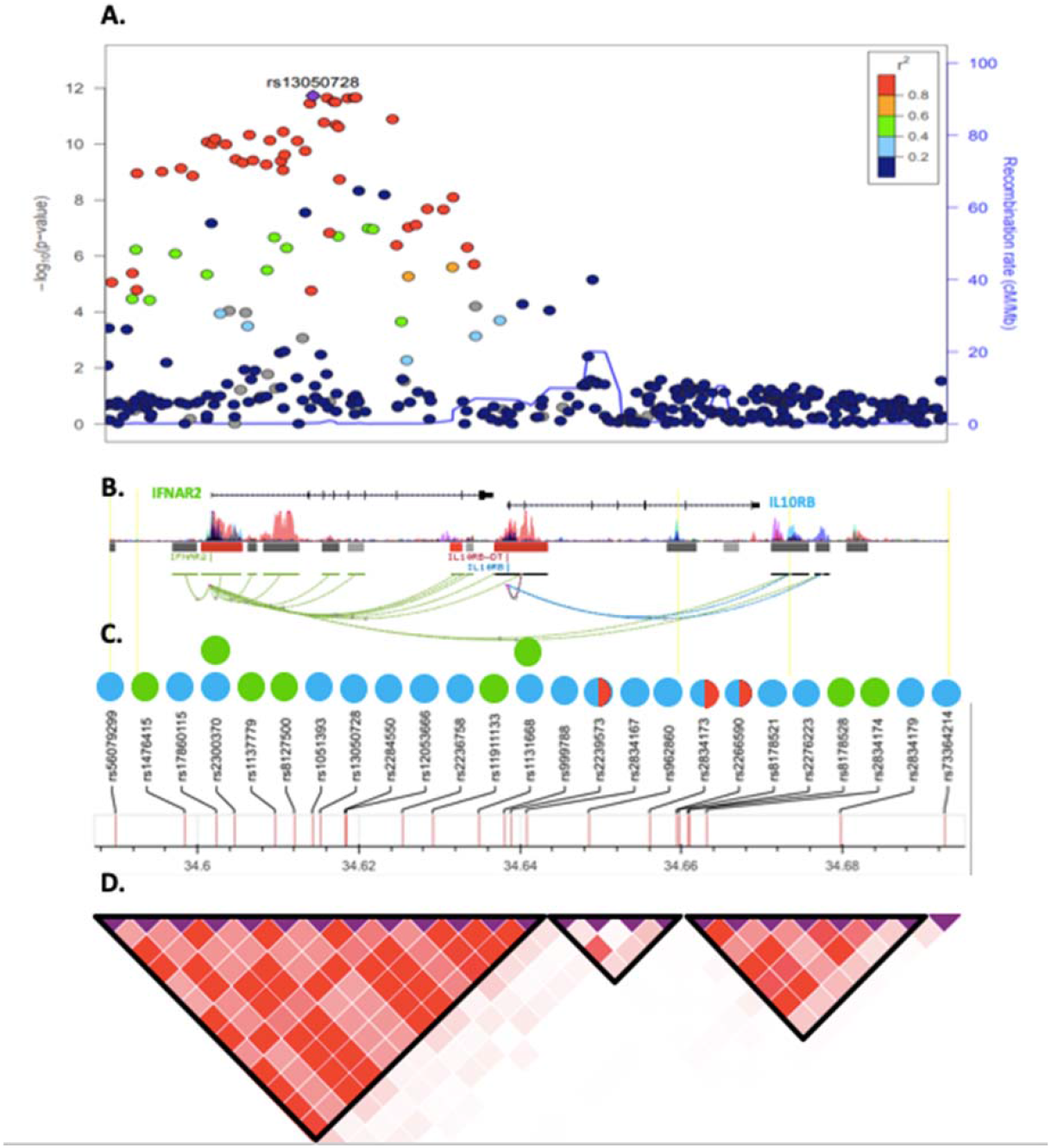
Genomic context, local association plot and LD structure of the *IFNAR2/IL10RB* region. **A**, Local association plot of the interval defined by all unique eQTLs for *IL10RB* or *IFNAR2*. Color code represents the degree of linkage disequilibrium with the most associated marker in 1000G Europeans. **B**, Genomic context of the region. Coding genes are represented by the refseq transcript. Bars represent epigenome Roadmap layered H3K27 acetylation markers. Connecting lines represent significant Hi-C interactions. **C**, Set of rsIDs used as proposed instruments for Mendelian Randomization analysis. Color code represents instruments for *IL10RB* (blue circles), *IFNAR2* (green circles). Red half-circles represent pQTLs for IL-10RB. **D** Linkage disequilibrium structure and blocks defined using European populations from 1000G project.

All significant MR results for *IFNAR2*/*IL10RB* that colocalized with COVID-19 hospitalization contained one of nine strongly correlated (*r*^2^>0.75 in 1000G European ancestry participants) variants (rs11911133, rs1051393, rs2300370, rs56079299, rs17860115, rs13050728, rs2236758, rs12053666, and rs1131668), which are *cis*-eQTLs for *IL10RB* in eleven tissues and for *IFNAR2* in four tissues (**Supplementary Table 8**). Within this LD block (hereafter rs13050728-LD block), rs13050728 is the eQTL most strongly associated with COVID-19 hospitalization (per T-allele odds ratio = 1.17; 95% CI = 1.12-1.23; *P*=1.88×10^−12^; **Supplementary Table 7**). Variants outside the rs13050728-LD block were not strongly associated with COVID-19 hospitalization (**Figure 3**).

#### pQTLs for IL10RB

Using stepwise conditional analysis on Olink measurements of plasma IL-10RB we identified two *cis*-pQTLs, rs2266590 (*P*=1.04×10^−136^) and rs2239573 (*P*= 2.66×10^−19^), which explained 5.4% and 1.2%, respectively, of the variance in plasma IL-10RB. rs2266590 was also an eQTL for *IL10RB* in three tissues and *IFNAR2* in one tissue, while rs2239573 was also an eQTL for *IL10RB* in two tissues (**Supplementary Table 8**). rs2266590 and rs2239573 lie in intron 5 and 1, respectively, of the *IL10RB* gene and are located in separate regions of high epigenetic modification (h3k27ac marking), indicating enhancer regions (**Figure 3**). rs2266590 and rs2239573 were not associated with COVID-19 hospitalization (*P*= 0.85 for rs2266590, *P*=0.66 for rs2239573, **Supplementary Figure 1**) and MR using these two *cis*-pQTLs yields a null result (*P*=0.74).

A third *cis*-pQTL (rs2834167, *P*=1.1×10^−8^) for plasma IL-10RB measured on the SomaScan platform was previously identified in 3,200 Icelanders over the age of 65.^18^ rs2834167 is a missense variant (Lys>Glu) and is not correlated with either of the *cis*-pQTLs for plasma IL-10RB measured by Olink (*r*^2^=0.01 for rs2266590, *r*^2^=0.03 for rs2239573 in 1000G EUR). Although rs2834167 was associated with *IL10RB* expression in 18 tissues, it was not associated with *IFNAR2* expression in any tissue (**Supplementary Table 8**). The A allele at rs2834167, which is associated with lower *IL10RB* gene expression but higher plasma IL-10RB, was inversely associated with COVID-19 (per-A-allele OR= 0.91; 95%CI= 0.87-0.95; *P*=5.3×10^−5^). Because Emilsson et al.^18^ did not report full summary statistics we could not perform colocalization between this pQTL and COVID-19 hospitalization. However, rs2834167 as an eQTL does not colocalize (PP.H4<0.8) with COVID-19 in any tissue (**Table 1**). These three *cis*-pQTLs, while possibly functional variants altering plasma IL-10RB levels, suggest that the plasma IL-10RB levels are not likely the mediator of the association between this locus and COVID-19 hospitalization. This suggests the MR assumption are unlikely to hold for IL10RB. IFNAR2 was not measured on the SomaScan or Olink platforms.

#### Phenome-wide scan of rs13050728

To identify other phenotypes associated with rs13050728, we performed a phenome-wide scan of publicly available data on PhenoScanner^19^ and GTEx, and unpublished proteomic data in INTERVAL (see methods). rs13050728 was associated with tryptase gamma 1 (TPSG1, *P*= 1.5×10^−5^) and vascular endothelial growth factor 2 (VEGFR2, *P*= 2.6×10^−5^, **Supplementary Table 9**), and both showed strong evidence of colocalization with COVID-19 hospitalization (PP.H4=0.96 for VEGFR2, PP.H4=0.96 for TPSG1, **Figure 4)**. The C allele at rs13050728 associated with higher *IFNAR2* expression in all tissues (except skeletal muscle), lower risk of COVID-19 hospitalization, and lower levels of plasma VEGFR2 and TPSG1 (**Supplementary Table 9**). This mimics agonistic effects of IFNAR2 through recombinant type-I IFNs, which are known to have an anti-angiogenic effect, at least in part through reduced VEGF/VEGFR2 signaling^20,21^, and decrease tryptase levels in a phase-2 trial using recombinant type-I IFN in patients with mastocytosis^22^, a condition that causes proliferation of mast cells. rs13050728 was not associated at *P*<4×10^−5^ (our Bonferroni corrected *P* value) with any phenotype beyond plasma VEGFR2 and TPSG1 and gene expression of *IFNAR2* and *IL10RB* (**Supplementary Table 9**), indicating that this variant is unlikely to exhibit widespread horizontal pleiotropy. Also, the chances of substantial bias due to MR violations is low^23^ since the variant is not strongly associated with other risk factors that could alter the likelihood of SARS-CoV-2 testing or hospitalization of COVID-19 patients.

**Figure 4.**
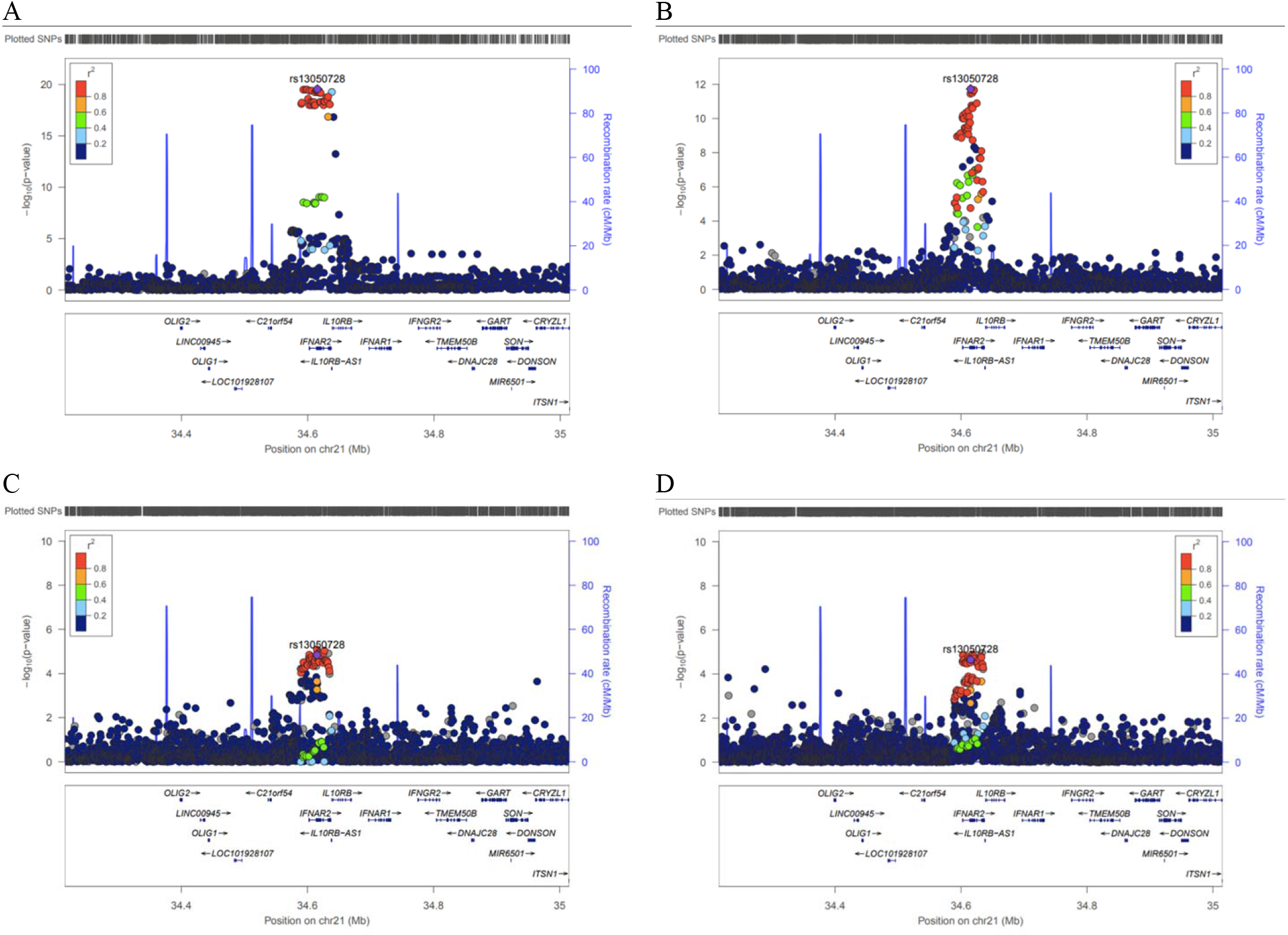
Regional association plots of the *IFNAR2*-*IL10RB* locus. Regional association plots for **A**, *IL10RB* gene expression in tibial nerve tissue from GTEx, **B** COVID-19 hospitalization from HGI and MVP trans-ancestry meta-analysis, **C, D**, Vascular endothelial growth factor receptor 2 (VEGFR2) and tryptase Gamma (TPSG1), respectively, measured by SomaLogic in 3,301 INTERVAL participants. All show the correlation (1000G European ancestry) for rs13050728, the *cis*-eQTL most associated with COVID-19 hospitalization in the *IL10RB*-I*FNAR2* region. All colocalize with each other (PP.H4>0.96 for all).

#### Pathway enrichment analysis of rs13050728

Using information from all GTEx V8 tissues we identified 476 genes whose expression levels were associated with rs13050728 at a nominal significance level (*P* < 0.05). Taking into consideration an adjusted *P* value for multiple testing within the WikiPathway corpus, only two biological pathways were significantly associated among all 624 pathways present in this database: Host-pathogen interaction of human corona viruses - IFN induction (adjusted *P* value = 0.0028) and Type I IFN Induction and Signaling During SARS-CoV-2 Infection (adjusted *P* value = 0.0098). In addition, among Gene Ontology (GO) and Reactome pathways, several gene sets were also significantly enriched. Notably, among enriched pathways were those related to IFN type I or antiviral response (**Supplementary Figure 2A**).

### ACE2

Angiotensin converting enzyme 2 (ACE2) converts angiotensin II into angiotensin (1-7) as part of the RAA system, and more importantly, is the viral receptor for SARS-CoV-2. We identified seven *cis*-eQTLs in seven tissues (**Supplementary Table 10**) for *ACE2* which are strongly correlated (*r*^2^>0.75 in 1000G EUR, **Supplementary Table 11**) with rs4830976 being the eQTL in the region most strongly associated with COVID-19 hospitalization.

#### pQTLs for ACE2

Stepwise conditional analysis for plasma ACE2 measured by Olink revealed one pQTL, rs5935998 (*P*=1.4×10^−21^), which is in high LD with a previously reported *cis*-pQTL (rs12558179) for ACE2 (*r*^2^=0.89 in 1000G EUR)^24^, and a secondary suggestive signal (rs4646156, *P*= 3.20×10^−7^). rs5935998 and rs4646156 are concordant in their effect on COVID-19 hospitalization (higher ACE2 levels corresponds to higher risk of COVID-19 hospitalization for both) resulting in a strong, positive MR association (MR beta-coefficient: 0.34; 95% CI: 0.17-0.51; *P*=8.1×10^−5^). Although neither rs5935998 or rs4646156 strongly colocalized with COVID-19 hospitalization (PP.H4=0.49 for rs5935998, PP.H4=0.08 for rs4646156, **Supplementary Figure 3**), the two pQTLs, while statistically independent, are mildly correlated (*r*^2^=0.2 in 1000G EUR), which can make colocalization difficult to interpret.^25^ One possible explanation is that these two pQTLs confer an effect on COVID-19 hospitalization that converges on the rs4830976-LD-block, as both are moderately correlated with rs4830976 (*r*^2^=0.32 for rs5935998, *r*^2^=0.42 for rs4646156 in 1000G EUR, **Supplementary Figure 3**)

#### Phenome-wide scan of rs4830976

rs4830976 is associated (*P*<4×10^−5^) with and colocalized (PP.H4>0.8) with expression of nearby genes *CA5B, CLTRN* (also known as *TMEM27*), and *VEGFD* (**Supplementary Table 12**) in at least one tissue, indicating that this variant may be instrumenting on gene expression beyond *ACE2*. However, given the biological prior that ACE2 acts as the receptor of SARS-CoV-2, *ACE2* is probably more likely than *CA5B, CLTRN* or *VEGFD* to be responsible for COVID-19 hospitalization. There were no other reported phenome-wide scan results at *P* <4×10^−5^ for rs4830976, which is at least in part due to the lack of reported X-chromosome results from a large proportion of GWAS.

#### Pathway enrichment analysis of rs4830976

Exploring the landscape of genes differentially expressed according to genotype in GTEx V8, we observed 1397 genes differentially expressed at a nominal *P* value less than 0.05. Over-representation analysis identified 238 significantly enriched biological pathways among differentially expressed genes (**Supplementary figure 2B**). Among these, signaling by interleukins, regulation of cytokine production, and antigen processing and presentation, might prove biologically relevant in COVID-19 infection.

## DISCUSSION

To identify drug-repurposing opportunities to inform trials against COVID-19, we conducted a large-scale MR analysis of protein and gene expression data. We first updated the “actionable” genome to an enlarged set of 1,263 human proteins and provided evidence for 700 of these as targets for drugs with some potential relevance to COVID-19. By testing more than a thousand of these using several of the largest currently available human genetic datasets, we provide evidence for drug targets of type-I IFNs (IFNAR2) and ACE2 modulators (ACE2) as priority candidates for evaluation in randomized trials of early management in COVID-19.

Our finding that ACE2 may play an important role in COVID-19 is unsurprising given its well-known relevance to SARS-CoV-2. Since ACE2 acts as the primary receptor for SARS-CoV-2, increased expression of ACE2 has been hypothesized to lead to increased susceptibility to infection. ACE2 plays a vital role in the RAAS signaling pathway, providing negative regulation through the conversion of Angiotensin II to Angiotensin 1-7. This action has anti-inflammatory and cardioprotective effects^26^ and plays a protective role in acute respiratory distress syndrome.^27,28^ ACE2 is a single-pass membrane protein but can be cleaved from the membrane to a soluble form which retains the enzymatic function to cleave Angiotensin II. It has therefore been hypothesized that administration of human recombinant soluble ACE2 (hrsACE2) could be an effective treatment for COVID-19, through distinct mechanisms in two phases of COVID-19. First, hrsACE2 can bind the viral spike glycoprotein of SARS-CoV-2, which could prevent cellular uptake of SARS-CoV-2 by reducing binding to the membrane-bound form of ACE2 (early phase). This suggestion is supported by the finding that APN01, a hrsACE2 therapeutic, showed a 1000-5000 fold reduction in SARS-CoV-2 viral load in primate kidney epithelial (Vero) cells, as well as inhibiting infection of human blood vessel and kidney organoids.^29^ In the later phase, hrsACE2 could reduce sequelae of SARS-CoV-2 infection by reducing inflammation in the lungs and other infected tissues. A case report of a hospitalized COVID-19 patient supported this hypothesis by showing that 7-day administration of APN01 was associated with a reduction in SARS-CoV-2 viral load and inflammatory markers.^30^ APN01 is currently being tested in a phase II trial to reduce mortality and invasive mechanical ventilation in 200 hospitalized COVID-19 patients^31^.

One of the main challenges of our analysis was to determine whether IFNAR2 or IL10RB (or both) was driving the association with COVID-19 hospitalization, given that they share *cis*-eQTLs used as proposed instruments for our MR analysis. Multiple lines of evidence indicate that IFNAR2 appears to be primarily responsible for the signal observed. First, our phenome-wide scan using the lead *IFNAR2/IL10RB cis*-eQTL reproduced known effects of type-I IFNs (the therapeutic target of IFNAR2) on VEGFR2 and TPSG1.^20-22^ Second, our pathway enrichment analysis using the same eQTL revealed pathways associated with type-I IFN receptor (IFNAR2) signaling. Last, three independent *cis*-pQTLs that are also *cis*-eQTLs for *IL10RB* did not show evidence of association with COVID-19, suggesting that plasma IL-10RB concentrations are unlikely to be etiologically relevant to COVID-19.

Evidence of a role for type-I IFN in COVID-19 is rapidly emerging. Studies using *in vitro* (A549 pulmonary cell lines), animal (ferrets) and *ex vivo* (human lung tissue) models have all shown lower expression of genes encoding type-I IFNs after exposure to SARS-CoV-2 compared to other respiratory viruses.^32,33^ This has been confirmed *in vivo* by studies showing significantly impaired type-I IFN response – including almost no IFN-beta activity - in the peripheral blood of severe COVID-19 patients compared to mild to moderate COVID-19 patients.^34^ More importantly, lower levels of IFN alpha-2 among recently hospitalized COVID-19 patients were associated with a substantial increase in the risk of progression to critical care, supporting our observation that lower genetically-predicted *IFNAR2* expression was associated with higher risk of COVID-19 hospitalization.^34^ Additionally, auto-antibodies for type 1 IFNs were found in a much higher proportion of individuals with severe COVID-19 than those with asymptomatic or mild SARS-CoV-2 infection.^35^

Whole exome and genome sequencing studies on severe COVID-19 patients have identified rare mutations that implicate type I IFN signaling. Zheng et al.^36^ found severe COVID-19 patients were enriched for rare variants predicted to cause loss of protein function at 13 genes involved in type-I IFN response. A cases-series of four severe COVID-19 patients under the age of 35 found a rare LOF mutation in *TLR7* and decreased type 1 IFN signaling.^37^

Several *in vitro* studies have shown that when diverse type of cells (including animal and human) and human organoids were pre-treated with type-I or -III IFNs, a reduction in SARS-CoV-2 replication was observed when compared with controls^38-42^ (**Supplementary Table 13**). Though these *in vitro* studies are encouraging, the evidence from randomized trials for type I IFNs in early COVID-19 stages is limited. Hung et al.^43^ showed that randomization to a combination of IFN beta-1b, ribavirin and lopinavir-ritonavir was superior to lopinavir-ritonavir alone in shortening the duration of viral shedding, alleviating symptoms and reducing the length of the hospital stay. Importantly, these benefits were confined to a subgroup who were hospitalized within 7 days of onset of symptoms where IFN beta-1b was administered to the intervention arm. These results, together with our genetic findings on COVID-19 hospitalization and the established role of type-I IFNs as first line of response against viral agents suggest recombinant type-I IFN as potential intervention during early stages of COVID-19. To date, there is no large randomized trial on IFN beta for early treatment of COVID-19 patients who are at high risk of hospitalization.

Trial evidence on the use of IFN-beta in late stages of COVID-19 has emerged in the last month. The SOLIDARITY trial, which randomized 2,050 hospitalized COVID-19 patients to IFN beta-1a, found no effect on mortality overall (relative risk (RR)=1.16, [95%CI: 0.96-1.39]), but possibly a trend across subgroups of COVID-19 severity at randomization (RR=1.40 [95%CI: 0.82-2.40] for those on ventilator, RR=1.13 [95%CI: 0.86-1.50] for those not ventilated but on oxygen, and RR=0.80 [95%CI: 0.27-2.35] in those with neither).^44^ The Adaptive COVID-19 Treatment Trial 3 (ACTT-3) trial stopped enrollment of severely ill COVID-19 patients for a trial on IFN beta-1a and remdesivir due to adverse events but continued enrolling patients with less severe disease.^45^ These findings indicate no role for the use of IFN beta during late stages of COVID-19, where the cytokine storm has already established.

Our study has several strengths. We provide an updated catalog of all actionable protein targets and drugs that are amenable to causal inference investigation through human genetics. By combining several of the most comprehensive datasets currently available and linking genetic variation with gene expression, plasma protein levels and COVID-19, we were able to robustly evaluate over one thousand actionable drug targets. We took multiple steps to minimize potential biases and confounding that could invalidate our actionable druggable-genome-wide MR analysis, such as using colocalization methods to minimize the chances of false positive results due to confounding by LD. We were careful to reduce the impact of horizontal pleiotropy by restricting our proposed instruments to variants acting in *cis* and performing phenome-wide scan to ensure they were only associated with gene expression of the tested gene or downstream phenotypes (vertical pleiotropy), but unmeasured sources can remain.

Our analysis also has limitations. Though we make use of instrumental variants from multiple data sources, none cover the entire actionable druggable genome, were ancestry-specific or were derived from COVID-19 patients. However, we managed to recover credible biological targets from our analysis that were consistent across ancestral groups. Identifying the most relevant tissue or cell-type can be challenging for interpreting MR analyses of gene expression. In our case, a relevant tissue could be: one invaded by SARS-CoV-2, an organ associated with clinical complications of COVID-19, a tissue where the COVID-19-relevant protein is produced, or a tissue that would be the likely site of action for the target drug. We opted to use a data-driven strategy that incorporates all tissues available in GTEx V8. For *IFNAR2*, we recovered fibroblasts (the main cell type responsible for IFN-beta production), esophageal mucosa^46,47^ (a tissue invaded by SARS-CoV-2), and skeletal muscle^48^ (associated with the neurological manifestations of COVID-19). For *ACE2*, we recovered brain tissue, an organ known to be invaded by SARS-CoV-2 and associated with clinical manifestations.^49,50^

In conclusion, our trans-ancestry MR analysis covering all actionable druggable genes identified two drug repurposing opportunities (type-I IFNs and hsrACE2) as interventions that need to be evaluated in adequately powered randomized trials to investigate their efficacy and safety for early management of COVID-19.

## METHODS

### Identification of actionable druggable genes suitable for repurposing against COVID-19

Information about drugs and clinical candidates, and their therapeutic targets, was obtained from the ChEMBL database (release 26^51^, **Supplementary Methods**). For the purposes of our COVID-19 drug repurposing efforts, actionable proteins were defined as those that are therapeutic targets of approved drugs and clinical candidates or are potential targets of approved drugs. Therapeutic targets were identified from the drug mechanism of action information in ChEMBL and linked to their component proteins. Each protein was assigned a confidence level based on the type and size of target annotated, and the resulting list was filtered to remove non-human proteins and those with lower confidence assignments (cases where the therapeutic target consists of more than 10 proteins or the protein is known to be a non-drug-binding subunit of a protein complex). For approved drugs, additional potential human target proteins were identified from pharmacological assay data in ChEMBL with recorded affinity/efficacy measurements <= 100nM (represented by a pChEMBL value >= 7).

A total of 1,263 unique human proteins were identified as ‘actionable’ from data available in ChEMBL. These consisted of 531 proteins that are therapeutic targets of approved drugs, 381 additional proteins that are therapeutic targets of clinical candidates and 351 additional proteins that are bound by approved drugs, but not annotated as the therapeutic targets. While the biological relevance of the latter group of targets in the context of the approved drug indications may be unclear, the high affinity/efficacy measurements suggest the drug should be capable of modulating these proteins, should they be found to be relevant to COVID-19 (although likely not in a selective manner). Proteins were further annotated with biological and drug information relating to their potential role in SARS-CoV-2 infection (**Supplementary methods**) such as change in abundance during infection, interaction with viral proteins or the activity of drugs in antiviral cell-based assays. Of the 1,263 actionable proteins identified previously, 300 were annotated as biologically relevant in SARS-CoV-2 infection and 547 were targets of drugs with some evidence of COVID-19 relevance from cell-based assays, clinical trials or the ATC classification (**Supplementary Table 2**).

### Selection of proposed instruments

#### eQTL proposed instruments

We proposed eQTL instruments using raw data from GTEx Version 8 by performing conditional analysis on normalized gene expression in European ancestry individuals in 49 tissues that had at least 70 samples. We used Matrix eQTL^52^ and followed the same procedure as outlined by the GTEx consortium (https://gtexportal.org/home/). Briefly, after filtering the genotypes (genotype missingness <0.05, MAF<0.01, HWE<0.000001, removing ambiguous SNPs), within each tissue, we performed GWAS between variants and gene expression adjusting for sex, the first 5 principal components of European genetic ancestry, PEER factors, sequencing platform and protocol. To identify independent eQTLs, we performed conditional analysis in regions around associations that fell below genome-wide significance, additionally adjusting for the peak variant if there exists an association reaching a *P*-value of 5×10^−8^. *Cis*-eQTLs were defined as significant (*P*<5×10^−8^) associations within 1Mb on either side of the encoded gene. To convert from build 38 to build 37, we used the table available from the GTEx consortium for all variants genotyped in GTEx v8 and hg19 liftover, (https://storage.googleapis.com/gtex_analysis_v8/reference/GTEx_Analysis_2017-06-05_v8_WholeGenomeSeq_838Indiv_Analysis_Freeze.lookup_table.txt.gz). In each tissue, multiple GW-significant (*P*<5×10^−8^) eQTLs for the same gene were combined into a single instrument. For example, for *IL10RB* expression in skeletal muscle tissue, there were two conditionally-independent eQTLs (rs2300370 and rs2834167, **Table 1**) that made up that instrument.

#### pQTL proposed instruments

We proposed pQTL instruments from two sources of publicly available data that reported conditionally independent pQTLs for proteins measured by the SomaLogic Inc. (Boulder, Colorado, US) SomaScan^53,54^ platform: (1) Sun *et al*.^15^, which reported results for 2,994 proteins in 3,301 INTERVAL participants and (2) Pietzner *et al*.^16^, which reported results for 179 proteins in 10,708 participants of the Fenland. In both, we restricted proposed instrumental variants to *cis*-pQTLs for actionable proteins, used a *P* value threshold of 5×10^−8^ and removed variants with MAF<0.01. MR was run independently for each data source (i.e. proposed instruments for the same protein in different platforms were tested against COVID-19 hospitalization independently).

### Estimates for COVID-19 hospitalization

To generate outcome summary-statistics, we meta-analyzed results from the Million Veteran Program (MVP), an ongoing, prospective cohort recruiting from 63 Veterans Health Administration (VA) medical facilities (**Supplementary Methods)**, and the Host Genetics Initiative,^10^ a global collaboration to accumulate GWAS on COVID-19 infection and clinical manifestations.

In MVP, 1,062 COVID-19 cases (**Supplementary Table 1)** were identified between March 1st and September 17, 2020 using an algorithm developed by the VA COVID National Surveillance Tool (NST). The NST classified COVID-19 cases as positive or negative based on reverse transcription polymerase chain reaction (rRT-PCR) laboratory test results conducted at VA clinics, supplemented with Natural Language Processing (NLP) on clinical documents. The algorithm to identify COVID-19 patients is continually updated to ensure new annotations of COVID-19 are captured from the clinical notes, with chart reviews performed periodically to validate the algorithm.^55^ COVID-19-related hospitalizations were defined as admissions from 7 days before up to 30 days after a patient’s first positive test for SARS-CoV-2 test. We tested association between all our proposed genetic instruments and COVID-19 hospitalization (versus population controls) in MVP adjusting for age, sex and the first 10 principal components in ancestry-specific strata using PLINK v2 (analysis completed on October 10, 2020). The MVP received ethical and study protocol approval by the Veterans Affairs Central Institutional Review Board and informed consent was obtained for all participants.

We downloaded publicly available summary statistics for the B2 outcome from Host Genetic Initiative on October 4, 2020 (release 4 version 1). In total, HGI accumulated 6,492 cases of COVID-19 hospitalization through collaboration from 16 contributing studies (**Supplementary Table 1**), which were asked to define cases as “hospitalized laboratory confirmed SARS-CoV-2 infection (RNA and/or serology based), hospitalization due to corona-related symptoms” versus population controls (https://docs.google.com/document/d/1okamrqYmJfa35ClLvCt_vEe4PkvrTwggHq7T3jbeyCI/view) and use a model that adjusts for age, age^2^, sex, age*sex, PCs, and study specific covariates (https://docs.google.com/document/d/16ethjgi4MzlQeO0KAW_yDYyUHdB9kKbtfuGW4XYVKQg/view) Results for each ancestry-stratum were meta-analyzed along with the HGI (summary statistics already meta-analyzed from contributing studies) for COVID hospitalization using METAL software^56^ with inverse-variance weighting and fixed effects.

### Mendelian randomization and colocalization

We conducted MR analyses using the R package TwoSampleMR (https://mrcieu.github.io/TwoSampleMR/). We used fixed-effects, inverse-variance weighted MR for proposed instruments that contain more than one variant, and Wald-ratio for proposed instruments with one variant. For proposed instruments with multiple variants, we also tested the heterogeneity across variant-level MR estimates, using the Cochrane Q method (mr_heterogeneity option in TwoSampleMR package). We defined significant MR results using a *P* value threshold of *P*<4.0×10^−5^ (0.05 Bonferroni-corrected for 1,263 actionable druggable genes) and identified a list of “suggestive” actionable druggable targets that passed a threshold of *P*<5×10^−4^. For statistically significant MR results, we also performed colocalization^57^ between each eQTL and the trans-ancestry meta-analysis on COVID-19 hospitalization using the moloc R package (https://github.com/clagiamba/moloc) with default priors (probability of shared causal variant for trait 1 and trait 2 is p1=p2=1×10^−4^, probability of shared causal variant across two traits is p12=1×10^−5^). For example, if a proposed instrument contained two variants, we performed colocalization for the primary eQTL GWAS with COVID-19 hospitalization, as well as the secondary eQTL GWAS (i.e. eQTL GWAS after adjusting for peak variant from primary GWAS) with COVID-19 hospitalization. Statistically significant MR hits with posterior probability for hypothesis-4 (PP.H4) > 0.8 (i.e. the probability of a shared causal variant) for a least one instrumental variant were then investigated further using the following analyses.

### Identifying pQTLs using Olink assay

We performed stepwise conditional analysis to identify *cis*-pQTL proposed instruments for proteins that passed our significance and colocalization thresholds and were one of 354 unique proteins measured on four Olink^58^ panels (CVD1, CVD2, Inflammation, and Neuro^59^) in 4,998 INTERVAL participants.^15^ INTERVAL is a prospective cohort study of ∼50,000 blood donors recruited from 25 National Health Service Blood and Transplant centers in England. Participants were genotyped using the UK Biobank Affymetrix Axiom array, followed by phasing using SHAPEIT3 and imputation on the Sanger Imputation Server using a 1000 Genomes Phase 3-UK10K imputation panel. Alleles were tested against Olink proteins using SNPTEST v2.5.2 and adjusted for age, sex, plate, time from blood draw to processing, season and the first 5 principal components. Conditional analysis was performed by adjusting for peak variants until no association fell below 5×10^−6^.

### Phenome-wide scan

We conducted a phenome-wide scan for variants with the following goals. First, we want to evaluate that our proposed instruments could reproduce the known phenotype associations (e.g. disease, biomarkers) ascribed to the drug that are due to on-target effects. Secondly, we want to identify if our proposed instruments are associated with comorbidities associated with greater likelihood of SARS-CoV-2 testing or predictors of hospitalization in COVID-19 patients, as this could potentially highlight the presence of certain biases.^23^ Also, for genes that were the target of licensed drugs, we checked whether the disease indication was also a risk factor for COVID-19 outcomes, as this might introduce a bias analogous to confounding by indication in MR.

To accomplish these goals, we investigated proposed instruments for associations of a phenome-wide range of outcomes. We searched the GTEx^14^ Portal (https://gtexportal.org/home/) for gene expression, and Phenoscanner^19^ (http://www.phenoscanner.medschl.cam.ac.uk/) for proteins, traits and diseases. We additionally queried variants in 354 Olink proteins data that have not been made publicly available (described earlier), and proteins measured by the SomaScan platform not previously published in Sun *et al*.^15^

### Characterizing downstream transcriptional consequences of associated loci

In order to confirm the specificity of the identified loci and to better explore their most important downstream transcriptional consequences, we have studied the transcriptional landscape modulation associated with the selected variants using GTEx V8 data with representation of 49 different tissues. For this we have used rs13050728 as the proxy of the *IFNAR2*/*IL10RB* locus and rs4830976 as the proxy of the *ACE2* locus and conducted a differential gene-expression analysis for all transcripts available in GTEx V8. After fitting models for all genes, enrichment pathway analysis was conducted to retrieve the most enriched pathways using both the differentially expressed (DE) gene list (through an over-representation analysis) and a Gene Set Enrichment Analysis framework (using the R package clusterProfiler^60^). For enrichment analysis we have used the corpus from WikiPathways, Gene Ontology and Reactome.

## Data Availability

Full MR results can be found on dbGAP in the VA Million Veteran Program space (accession: phs001672).

## Author contribution

J.P.C., A.S.B. and J.M.G. conceived the study design. A.G., A.P.B. and A.R.L. defined the actionable genome, and identified and curated drug information relating to SARS-CoV-2; P.B. and I.B.-H. provided biological annotation relating to SARS-CoV-2; C.G. performed stepwise conditional analysis on GTEx raw data; D.P. tested associations for COVID-19 in MVP; L.G. and J.H.Z. performed meta-analysis of HGI and MVP; L.G. performed Mendelian randomization analysis; L.G. and C.G. performed colocalization analyses; L.G. and B.P.P. performed conditional analysis on Olink proteins; L.G. and E.A. performed phenome-wide scans; A.C.P. performed pathway enrichment analysis; J.N.D., A.S.B., and J.E.P. provided INTERVAL data; Several authors were involved in the curation of the MVP data; L.G., C.G., A.C.P., A.G., D.P., A.S.B. and J.P.C. wrote the manuscript. J.P.C. oversaw all analyses. All authors critically reviewed the manuscript.

## Acknowledgements

We are grateful to the Host Genetic Initiative for making their data publicly available (full acknowledgements can be found here: https://www.covid19hg.org/acknowledgements/). This research is based on data from the Million Veteran Program, Office of Research and Development, Veterans Health Administration, and was supported by award #MVP035. This research was also supported by additional Department of Veterans Affairs awards grant #MVP001. This publication does not represent the views of the Department of Veteran Affairs or the United States Government. Full acknowledgements for the VA Million Veteran Program COVID-19 Science Initiative can be found in the supplementary methods. C.G. has received funding from the European Union’s Horizon 2020 research and innovation program under the Marie Sklodowska-Curie grant agreement No 754490 – MINDED project. A.G., P.B. and A.R.L. are funded by the Member States of the European Molecular Biology Laboratory (EMBL). I.B.- H. received funding from Open Targets (grant agreement OTAR-044). The Fenland Study (10.22025/2017.10.101.00001) is funded by the Medical Research Council (MC_UU_12015/1); we are grateful to all the volunteers and to the General Practitioners and practice staff for assistance with recruitment; we thank the Fenland Study Investigators, Fenland Study Co-ordination team and the Epidemiology Field, Data and Laboratory teams; we further acknowledge support for genomics from the Medical Research Council (MC_PC_13046); proteomic measurements were supported and governed by a collaboration agreement between the University of Cambridge and Somalogic. J.E.P. is supported by UKRI Innovation Fellowship at Health Data Research UK (MR/S004068/2). L.R., N.H. and C.L. are supported by the Swedish Research Council.

