## Supplementary Methods for "Actionable druggable genome-wide Mendelian randomization identifies repurposing opportunities for COVID-19"

#### CHEMBL Database

ChEMBL curates approved drug information from the FDA Orange Book, WHO Anatomical Therapeutic Chemical classification and British National Formulary and clinical candidate information from ClinicalTrials.gov, United States Adopted Name applications and pharmaceutical company pipeline documents. The therapeutic targets of these compounds are manually annotated using a variety of sources including peer-reviewed literature and drug prescribing information, focusing on those proteins with which the drug directly interacts and that are believed to be responsible for the efficacy of the drug in the approved or clinically tested indications[1]. As well as individual proteins, targets in ChEMBL can represent protein complexes, protein families and other molecular groups that drugs may interact with and can therefore consist of multiple proteins. ChEMBL also includes extensive pharmacology data for drugs, candidates and preclinical/research compounds, extracted from Medicinal Chemistry literature. Such data can give supplementary information about additional proteins with which drugs may interact that could be responsible for adverse effects or have unknown biological relevance.

#### COVID-19 relevant drugs

Nine publications and preprints were identified that contained cell-based screening data of drugs and clinical candidates against SARS-CoV-2.[2-10] Drug names were extracted from each publication and mapped to ChEMBL compound identifiers[11], using a combination of automated mapping and manual curation, in order to align data from different publications. Drugs were considered active if they either had IC50 or EC50 measurements <10uM or had other measurements types and were defined as active by the authors.

Drugs and biologics being tested in interventional clinical trials for COVID-19 were identified from the ClinicalTrials.gov database (downloaded 07/20/20). Drug names were mapped to ChEMBL compound identifiers, where possible, and the phase of each trial captured. Blood products, cell-based therapies, vaccines and natural product mixtures were excluded.

Approved drugs with anti-inflammatory or immunomodulatory action were identified from ChEMBL (release 26) using ATC codes with the following prefixes: L03, L04, M01, A07E, H02, R06, R03BA and R01AD. Approved drugs with anti-coagulant action were also identified using ATC codes with the B01A prefix. COVID-relevant drugs were then mapped to the list of actionable genes via the curated therapeutic targets and potential target information in ChEMBL.

**Biological relevance to SARS-CoV-2 infection**

We further annotated proteins that have biological evidence relating to their potential role in SARS-CoV-2 infection. This information included whether the abundance of the protein is regulated after SARS-CoV-2 infection[12], whether the protein is believed to interact with viral proteins from SARS-CoV-2, SARS-CoV-1 or MERS-CoV[13], whether the protein is implicated in other viral infections (https://covid19.opentargets.org/), the number of publications mentioning COVID-19 and the protein in the same sentence (https://covid19.opentargets.org/), information concerning human phosphoproteome changes during infection with SARS-CoV-2 (phosphosite dynamics as well as kinase activities)[10], and finally, data regarding the effect of KO/KD of proteins during infection with SARS-CoV2 (CRISPR in Caco2 cells and iRNA in A549).[13]

**Safety information**

Information about potential safety issues associated with proteins was extracted from the Open Targets platform.[14] This information includes known target safety risks identified from the literature and the HeCaTos projects, and experimentally identified safety risks from the eTox and TOX21 data sets. Safety issues relating to specific drugs were also collated from ChEMBL. This included whether any products containing the drug carry a boxed warning on their package label, whether the drug has been withdrawn for safety or efficacy reasons in at least one country, and finally whether the drug showed significant cytotoxicity in any of the cell-based SARS-CoV-2 screens described above (defined as a selectivity index <= 2).

### Populations

#### *Million Veteran Program*

MVP is an ongoing, prospective biobank that recruits from 63 Veterans Health Administration (VA) medical facilities. Participants are linked to VA electronic health records (EHR), complete a questionnaire, and submit a blood sample at enrollment. Genotyping was performed using the MVP array, which is based on the UK Biobank Affymetrix Axiom array but with modifications tailored to the veteran population.[15] Samples were phased using EAGLE v2 software[16] and imputed with Minimac3[17] using the 1000 Genomes Project reference panel (phase 3, version 5).[18] Ancestry was determined through the HARE (harmonized ancestry and race/ethnicity) approach, which combines self-identified race/ethnicity and genetic information to more accurately determine ancestry groups.[19] The Veterans Affairs (VA) central institutional review board (IRB) and site-specific Research and Development (R&D) committees approved the Million Veteran Program study.

**VA Million Veteran Program COVID-19 Science Initiative:**
**Core Acknowledgement for Publications**
**Updated November 17, 2020**

### MVP COVID-19 Science Program Steering Committee

- Christopher J. O’Donnell, M.D., M.P.H. (Co-Chair)
  VA Boston Healthcare System, 150 S. Huntington Avenue, Boston, MA 02130
- J. Michael Gaziano, M.D., M.P.H. (Co-Chair)
  VA Boston Healthcare System, 150 S. Huntington Avenue, Boston, MA 02130
- Philip S. Tsao, Ph.D. (Co-Chair)
  VA Palo Alto Health Care System, 3801 Miranda Avenue, Palo Alto, CA 94304
- Sumitra Muralidhar, Ph.D.
  US Department of Veterans Affairs, 810 Vermont Avenue NW, Washington, DC 20420
- Jean Beckham, Ph.D.
  Durham VA Medical Center, 508 Fulton Street, Durham, NC 27705
- Kyong-Mi Chang, M.D.
  Philadelphia VA Medical Center, 3900 Woodland Avenue, Philadelphia, PA 19104
- JP Casas Romero, M.D., Ph.D.
  VA Boston Healthcare System, 150 S. Huntington Avenue, Boston, MA 02130
- Kelly Cho, M.P.H., Ph.D.
  VA Boston Healthcare System, 150 S. Huntington Avenue, Boston, MA 02130
- Saiju Pyarajan, Ph.D.
  VA Boston Healthcare System, 150 S. Huntington Avenue, Boston, MA 02130
- Jennifer Huffman, Ph.D.
  VA Boston Healthcare System, 150 S. Huntington Avenue, Boston, MA 02130

### MVP COVID-19 Science Program Steering Committee Support

- Lauren Thomann, M.P.H. (P&P Committee Representative, Working Group Coordinator)
  VA Boston Healthcare System, 150 S. Huntington Avenue, Boston, MA 02130
- Helene Garcon, M.D. (Program Coordinator, Working Group Coordinator)
  VA Boston Healthcare System, 150 S. Huntington Avenue, Boston, MA 02130
- Nicole Kosik, M.P.H. (Working Group Coordinator)
  VA Boston Healthcare System, 150 S. Huntington Avenue, Boston, MA 02130

### MVP COVID-19 Science Program Working Groups and Associated Chairs

- COVID-19 Related PheWAS
  - Katherine Liao, M.D.
    VA Boston Healthcare System, 150 S. Huntington Avenue, Boston, MA 02130
  - Scott Damrauer, M.D.
    Philadelphia VA Medical Center, 3900 Woodland Avenue, Philadelphia, PA 19104

- Disease Mechanisms
    - Richard Hauger, M.D.
      VA San Diego Healthcare System, 3350 La Jolla Village Drive, San Diego, CA 92161
    - Shiuh-Wen Luoh, M.D., Ph.D.
      Portland VA Medical Center, 3710 SW U.S. Veterans Hospital Road, Portland, OR 97239
    - Sudha Iyengar, Ph.D.
      VA Northeast Ohio Healthcare System, 10701 East Boulevard, Cleveland, OH 44106
- Druggable Genome
    - JP Casas Romero, M.D., Ph.D.
      VA Boston Healthcare System, 150 S. Huntington Avenue, Boston, MA 02130
    - Todd Edwards, Ph.D.
      VA Tennessee Valley Healthcare System, 2525 West End Avenue, Nashville, TN 37204
- Genomics for Risk Prediction, PRS, and MR
    - Themistocles Assimes, M.D., Ph.D.
      VA Palo Alto Health Care System, 3801 Miranda Avenue, Palo Alto, CA 94304
    - Panagiotis Roussos, M.D., Ph.D.
      James J. Peters VA Medical Center, 130 W Kingsbridge Rd, Bronx, NY 10468
    - Robert Striker, M.D., Ph.D.
      William S. Middleton Memorial Veterans Hospital, 2500 Overlook Terrace, Madison, WI 53705
- GWAS & Downstream Analysis
    - Jennifer Huffman, Ph.D.
      VA Boston Healthcare System, 150 S. Huntington Avenue, Boston, MA 02130
    - Yan Sun, Ph.D.
      Atlanta VA Medical Center, 1670 Clairmont Road, Decatur, GA 30033
- Pharmacogenomics
    - Adriana Hung, M.D., M.P.H.
      VA Tennessee Valley Healthcare System, 1310 24th Avenue, South Nashville, TN 37212
    - Sony Tuteja, Pharm.D., M.S.
      Philadelphia VA Medical Center, 3900 Woodland Avenue, Philadelphia, PA 19104
- VA COVID-19 Shared Data Resource – Scott L. DuVall, Ph.D.; Kristine E. Lynch, Ph.D.; Elise Gatsby, M.P.H.
  VA Informatics and Computing Infrastructure (VINCI), VA Salt Lake City Health Care System, 500 Foothill Drive, Salt Lake City, UT 84148

- MVP COVID-19 Data Core – Kelly Cho, M.P.H., Ph.D.; Lauren Costa, M.P.H.; Anne Yuk-Lam Ho, M.P.H.; Rebecca Song, M.P.H.
  VA Boston Healthcare System, 150 S. Huntington Avenue, Boston, MA 02130

**MVP Executive Committee**

- Co-Chair: J. Michael Gaziano, M.D., M.P.H.
  VA Boston Healthcare System, 150 S. Huntington Avenue, Boston, MA 02130
- Co-Chair: Sumitra Muralidhar, Ph.D.
  US Department of Veterans Affairs, 810 Vermont Avenue NW, Washington, DC 20420
- Rachel Ramoni, D.M.D., Sc.D., Chief VA Research and Development Officer
  US Department of Veterans Affairs, 810 Vermont Avenue NW, Washington, DC 20420
- Jean Beckham, Ph.D.
  Durham VA Medical Center, 508 Fulton Street, Durham, NC 27705
- Kyong-Mi Chang, M.D.
  Philadelphia VA Medical Center, 3900 Woodland Avenue, Philadelphia, PA 19104
- Christopher J. O'Donnell, M.D., M.P.H.
  VA Boston Healthcare System, 150 S. Huntington Avenue, Boston, MA 02130
- Philip S. Tsao, Ph.D.
  VA Palo Alto Health Care System, 3801 Miranda Avenue, Palo Alto, CA 94304
- James Breeling, M.D., Ex-Officio
  US Department of Veterans Affairs, 810 Vermont Avenue NW, Washington, DC 20420
- Grant Huang, Ph.D., Ex-Officio
  US Department of Veterans Affairs, 810 Vermont Avenue NW, Washington, DC 20420
- JP Casas Romero, M.D., Ph.D., Ex-Officio
  VA Boston Healthcare System, 150 S. Huntington Avenue, Boston, MA 02130

**MVP Program Office**

- Sumitra Muralidhar, Ph.D.
  US Department of Veterans Affairs, 810 Vermont Avenue NW, Washington, DC 20420
- Jennifer Moser, Ph.D.
  US Department of Veterans Affairs, 810 Vermont Avenue NW, Washington, DC 20420

**MVP Recruitment/Enrollment**

- Recruitment/Enrollment Director/Deputy Director, Boston – Stacey B. Whitbourne, Ph.D.; Jessica V. Brewer, M.P.H.
  VA Boston Healthcare System, 150 S. Huntington Avenue, Boston, MA 02130
- MVP Coordinating Centers
  - Clinical Epidemiology Research Center (CERC), West Haven – Mihaela Aslan, Ph.D.
    West Haven VA Medical Center, 950 Campbell Avenue, West Haven, CT 06516
  - Cooperative Studies Program Clinical Research Pharmacy Coordinating Center, Albuquerque – Todd Connor, Pharm.D.; Dean P. Argyres, B.S., M.S.

New Mexico VA Health Care System, 1501 San Pedro Drive SE, Albuquerque, NM 87108

  - Genomics Coordinating Center, Palo Alto – Philip S. Tsao, Ph.D.
    VA Palo Alto Health Care System, 3801 Miranda Avenue, Palo Alto, CA 94304
  - MVP Boston Coordinating Center, Boston - J. Michael Gaziano, M.D., M.P.H.
    VA Boston Healthcare System, 150 S. Huntington Avenue, Boston, MA 02130
  - MVP Information Center, Canandaigua – Brady Stephens, M.S.
    Canandaigua VA Medical Center, 400 Fort Hill Avenue, Canandaigua, NY 14424

- VA Central Biorepository, Boston – Mary T. Brophy M.D., M.P.H.; Donald E. Humphries, Ph.D.; Luis E. Selva, Ph.D.
  VA Boston Healthcare System, 150 S. Huntington Avenue, Boston, MA 02130
- MVP Informatics, Boston – Nhan Do, M.D.; Shahpoor (Alex) Shayan, M.S.
  VA Boston Healthcare System, 150 S. Huntington Avenue, Boston, MA 02130
- MVP Data Operations/Analytics, Boston – Kelly Cho, M.P.H., Ph.D.
  VA Boston Healthcare System, 150 S. Huntington Avenue, Boston, MA 02130
- Director of Regulatory Affairs – Lori Churby, B.S.
  VA Palo Alto Health Care System, 3801 Miranda Avenue, Palo Alto, CA 94304

**MVP Science**

- Science Operations – Christopher J. O'Donnell, M.D., M.P.H.
  VA Boston Healthcare System, 150 S. Huntington Avenue, Boston, MA 02130
- Genomics Core - Christopher J. O'Donnell, M.D., M.P.H.
  VA Boston Healthcare System, 150 S. Huntington Avenue, Boston, MA 02130
  Saiju Pyarajan Ph.D.
  VA Boston Healthcare System, 150 S. Huntington Avenue, Boston, MA 02130
  Philip S. Tsao, Ph.D.
  VA Palo Alto Health Care System, 3801 Miranda Avenue, Palo Alto, CA 94304
- Data Core - Kelly Cho, M.P.H, Ph.D.
  VA Boston Healthcare System, 150 S. Huntington Avenue, Boston, MA 02130
- VA Informatics and Computing Infrastructure (VINCI) – Scott L. DuVall, Ph.D.
  VA Salt Lake City Health Care System, 500 Foothill Drive, Salt Lake City, UT 84148
- Data and Computational Sciences – Saiju Pyarajan, Ph.D.
  VA Boston Healthcare System, 150 S. Huntington Avenue, Boston, MA 02130
- Statistical Genetics – Elizabeth Hauser, Ph.D.
  Durham VA Medical Center, 508 Fulton Street, Durham, NC 27705
  Yan Sun, Ph.D.
  Atlanta VA Medical Center, 1670 Clairmont Road, Decatur, GA 30033
  Hongyu Zhao, Ph.D.
  West Haven VA Medical Center, 950 Campbell Avenue, West Haven, CT 06516

**Current MVP Local Site Investigators**

- Atlanta VA Medical Center (Peter Wilson, M.D.)
  1670 Clairmont Road, Decatur, GA 30033
- Bay Pines VA Healthcare System (Rachel McArdle, Ph.D.)
  10,000 Bay Pines Blvd Bay Pines, FL 33744
- Birmingham VA Medical Center (Louis Dellitalia, M.D.)
  700 S. 19th Street, Birmingham AL 35233
- Central Western Massachusetts Healthcare System (Kristin Mattocks, Ph.D., M.P.H.)
  421 North Main Street, Leeds, MA 01053
- Cincinnati VA Medical Center (John Harley, M.D., Ph.D.)
  3200 Vine Street, Cincinnati, OH 45220
- Clement J. Zablocki VA Medical Center (Jeffrey Whittle, M.D., M.P.H.)
  5000 West National Avenue, Milwaukee, WI 53295
- VA Northeast Ohio Healthcare System (Frank Jacono, M.D.)
  10701 East Boulevard, Cleveland, OH 44106
- Durham VA Medical Center (Jean Beckham, Ph.D.)
  508 Fulton Street, Durham, NC 27705
- Edith Nourse Rogers Memorial Veterans Hospital (John Wells., Ph.D.)
  200 Springs Road, Bedford, MA 01730
- Edward Hines, Jr. VA Medical Center (Salvador Gutierrez, M.D.)
  5000 South 5th Avenue, Hines, IL 60141
- Veterans Health Care System of the Ozarks (Gretchen Gibson, D.D.S., M.P.H.)
  1100 North College Avenue, Fayetteville, AR 72703
- Fargo VA Health Care System (Kimberly Hammer, Ph.D.)
  2101 N. Elm, Fargo, ND 58102
- VA Health Care Upstate New York (Laurence Kaminsky, Ph.D.)
  113 Holland Avenue, Albany, NY 12208
- New Mexico VA Health Care System (Gerardo Villareal, M.D.)
  1501 San Pedro Drive, S.E. Albuquerque, NM 87108
- VA Boston Healthcare System (Scott Kinlay, M.B.B.S., Ph.D.)
  150 S. Huntington Avenue, Boston, MA 02130
- VA Western New York Healthcare System (Junzhe Xu, M.D.)
  3495 Bailey Avenue, Buffalo, NY 14215-1199
- Ralph H. Johnson VA Medical Center (Mark Hamner, M.D.)
  109 Bee Street, Mental Health Research, Charleston, SC 29401
- Columbia VA Health Care System (Roy Mathew, M.D.)
  6439 Garners Ferry Road, Columbia, SC 29209
- VA North Texas Health Care System (Sujata Bhushan, M.D.)
  4500 S. Lancaster Road, Dallas, TX 75216
- Hampton VA Medical Center (Pran Iruvanti, D.O., Ph.D.)
  100 Emancipation Drive, Hampton, VA 23667

- Richmond VA Medical Center (Michael Godschalk, M.D.)
  1201 Broad Rock Blvd., Richmond, VA 23249
- Iowa City VA Health Care System (Zuhair Ballas, M.D.)
  601 Highway 6 West, Iowa City, IA 52246-2208
- Eastern Oklahoma VA Health Care System (Douglas Ivins, M.D.)
  1011 Honor Heights Drive, Muskogee, OK 74401
- James A. Haley Veterans' Hospital (Stephen Mastorides, M.D.)
  13000 Bruce B. Downs Blvd, Tampa, FL 33612
- James H. Quillen VA Medical Center (Jonathan Moorman, M.D., Ph.D.)
  Corner of Lamont & Veterans Way, Mountain Home, TN 37684
- John D. Dingell VA Medical Center (Saib Gappy, M.D.)
  4646 John R Street, Detroit, MI 48201
- Louisville VA Medical Center (Jon Klein, M.D., Ph.D.)
  800 Zorn Avenue, Louisville, KY 40206
- Manchester VA Medical Center (Nora Ratcliffe, M.D.)
  718 Smyth Road, Manchester, NH 03104
- Miami VA Health Care System (Hermes Florez, M.D., Ph.D.)
  1201 NW 16th Street, 11 GRC, Miami FL 33125
- Michael E. DeBakey VA Medical Center (Olaoluwa Okusaga, M.D.)
  2002 Holcombe Blvd, Houston, TX 77030
- Minneapolis VA Health Care System (Maureen Murdoch, M.D., M.P.H.)
  One Veterans Drive, Minneapolis, MN 55417
- N. FL/S. GA Veterans Health System (Peruvemba Sriram, M.D.)
  1601 SW Archer Road, Gainesville, FL 32608
- Northport VA Medical Center (Shing Shing Yeh, Ph.D., M.D.)
  79 Middleville Road, Northport, NY 11768
- Overton Brooks VA Medical Center (Neeraj Tandon, M.D.)
  510 East Stoner Ave, Shreveport, LA 71101
- Philadelphia VA Medical Center (Darshana Jhala, M.D.)
  3900 Woodland Avenue, Philadelphia, PA 19104
- Phoenix VA Health Care System (Samuel Aguayo, M.D.)
  650 E. Indian School Road, Phoenix, AZ 85012
- Portland VA Medical Center (David Cohen, M.D.)
  3710 SW U.S. Veterans Hospital Road, Portland, OR 97239
- Providence VA Medical Center (Satish Sharma, M.D.)
  830 Chalkstone Avenue, Providence, RI 02908
- Richard Roudebush VA Medical Center (Suthat Liangpunsakul, M.D., M.P.H.)
  1481 West 10th Street, Indianapolis, IN 46202
- Salem VA Medical Center (Kris Ann Oursler, M.D.)
  1970 Roanoke Blvd, Salem, VA 24153

- San Francisco VA Health Care System (Mary Whooley, M.D.)
  4150 Clement Street, San Francisco, CA 94121
- South Texas Veterans Health Care System (Sunil Ahuja, M.D.)
  7400 Merton Minter Boulevard, San Antonio, TX 78229
- Southeast Louisiana Veterans Health Care System (Joseph Constans, Ph.D.)
  2400 Canal Street, New Orleans, LA 70119
- Southern Arizona VA Health Care System (Paul Meyer, M.D., Ph.D.)
  3601 S 6th Avenue, Tucson, AZ 85723
- Sioux Falls VA Health Care System (Jennifer Greco, M.D.)
  2501 W 22nd Street, Sioux Falls, SD 57105
- St. Louis VA Health Care System (Michael Rauchman, M.D.)
  915 North Grand Blvd, St. Louis, MO 63106
- Syracuse VA Medical Center (Richard Servatius, Ph.D.)
  800 Irving Avenue, Syracuse, NY 13210
- VA Eastern Kansas Health Care System (Melinda Gaddy, Ph.D.)
  4101 S 4th Street Trafficway, Leavenworth, KS 66048
- VA Greater Los Angeles Health Care System (Agnes Wallbom, M.D., M.S.)
  11301 Wilshire Blvd, Los Angeles, CA 90073
- VA Long Beach Healthcare System (Timothy Morgan, M.D.)
  5901 East 7th Street Long Beach, CA 90822
- VA Maine Healthcare System (Todd Stapley, D.O.)
  1 VA Center, Augusta, ME 04330
- VA New York Harbor Healthcare System (Scott Sherman, M.D., M.P.H.)
  423 East 23rd Street, New York, NY 10010
- VA Pacific Islands Health Care System (George Ross, M.D.)
  459 Patterson Rd, Honolulu, HI 96819
- VA Palo Alto Health Care System (Philip Tsao, Ph.D.)
  3801 Miranda Avenue, Palo Alto, CA 94304-1290
- VA Pittsburgh Health Care System (Patrick Strollo, Jr., M.D.)
  University Drive, Pittsburgh, PA 15240
- VA Puget Sound Health Care System (Edward Boyko, M.D.)
  1660 S. Columbian Way, Seattle, WA 98108-1597
- VA Salt Lake City Health Care System (Laurence Meyer, M.D., Ph.D.)
  500 Foothill Drive, Salt Lake City, UT 84148
- VA San Diego Healthcare System (Samir Gupta, M.D., M.S.C.S.)
  3350 La Jolla Village Drive, San Diego, CA 92161
- VA Sierra Nevada Health Care System (Mostaqul Huq, Pharm.D., Ph.D.)
  975 Kirman Avenue, Reno, NV 89502
- VA Southern Nevada Healthcare System (Joseph Fayad, M.D.)
  6900 North Pecos Road, North Las Vegas, NV 89086

- VA Tennessee Valley Healthcare System (Adriana Hung, M.D., M.P.H.)
  1310 24th Avenue, South Nashville, TN 37212
- Washington DC VA Medical Center (Jack Lichy, M.D., Ph.D.)
  50 Irving St, Washington, D. C. 20422
- W.G. (Bill) Hefner VA Medical Center (Robin Hurley, M.D.)
  1601 Brenner Ave, Salisbury, NC 28144
- White River Junction VA Medical Center (Brooks Robey, M.D.)
  163 Veterans Drive, White River Junction, VT 05009
- William S. Middleton Memorial Veterans Hospital (Robert Striker, M.D., Ph.D.)
  2500 Overlook Terrace, Madison, WI 53705
