## Supplementary Figures for "Actionable druggable genome-wide Mendelian randomization identifies repurposing opportunities for COVID-19"

**Supplementary Figure 1.** Regional association plots for rs2266590 and rs2239573 for plasma IL-10RB, *IL10RB* expression, and COVID-19 hospitalization. **A,** rs2266590 as pQTL for plasma IL10RB, Olink INTERVAL. **B,** rs2266590 as eQTL for *IL10RB* expression (after adjusting for rs2834167 and rs13050728) in tibial nerve tissue. **C,** rs2266590 in COVID-19 hospitalization. **D,** rs2239573 as pQTL for plasma IL-10RB, Olink INTERVAL (after adjusting for rs2266590). **E,** rs2239573 as eQTL for IL10RB expression in whole blood. **F,** rs2239573 in COVID-19 hospitalization. **A** colocalizes with **B**, and **D** colocalizes with **E**. These two variants, that are *cis*-eQTLs and *cis*-pQTLs for IL10RB, are not associated with COVID-19 hospitalization ($p = 0.85$ for rs2266590, $p=0.66$ for rs2239573).

**Supplementary Figure 2.** Enrichment analysis of peak eQTLs for *IFNAR2-IL10RB* and *ACE2* regions. Results obtained from association analysis using all 49 tissues from GTEx V8 contrasted against variant genotypes in an additive model. Dotplot of over-representation analysis using all significant ($p<0.05$) differentially expressed (DE) genes (476 for rs13050728; 1,397 for rs4830976) for **A,** rs13050728, peak eQTL in the *IFNAR2-IL10RB* region and **B,** rs4830976, peak eQTL in the *ACE2* region. Count = number of DE genes part of the enriched pathway. Gene ratio is the rate of DE genes represented in each pathway.

**Supplementary Figure 3.** Regional association plots for *ACE2* expression, and their association with COVID-19 hospitalization. **A,** rs4830976 for *ACE2* expression in brain frontal cortex tissue. **B,** rs4830976 in COVID-19 hospitalization. **C,** primary pQTL (rs5935998) for plasma ACE2 measured by Oink in 4,994 INTERVAL participants **D,** rs5935998 in COVID-19 hospitalization **E,** secondary pQTL (rs4646156) for plasma ACE2 measured by Oink in 4,994 INTERVAL participants. **F,** rs4646156 in COVID-19 hospitalization.

**Supplementary Figure 1.**

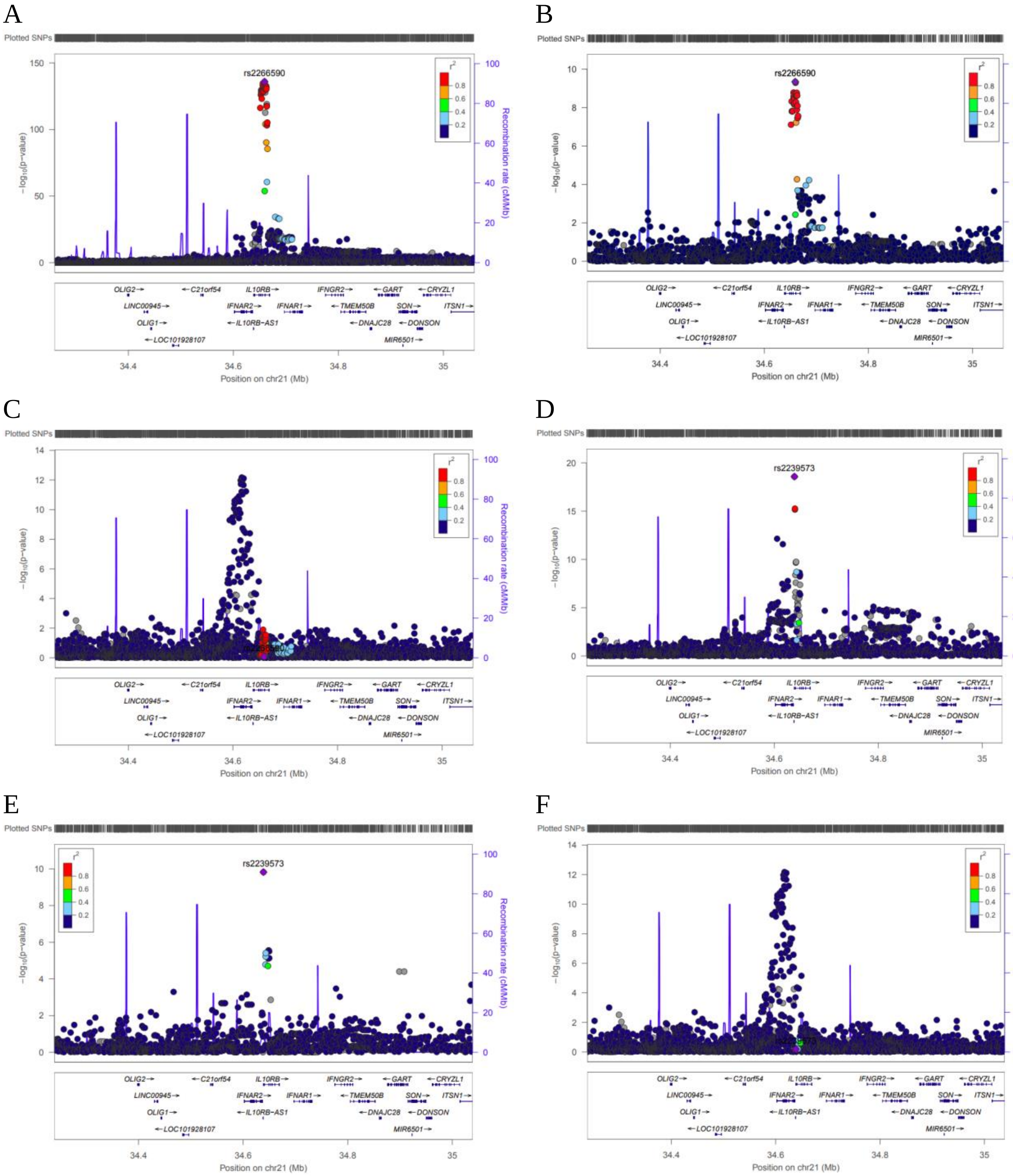

**Supplementary figure 2.**

**A**

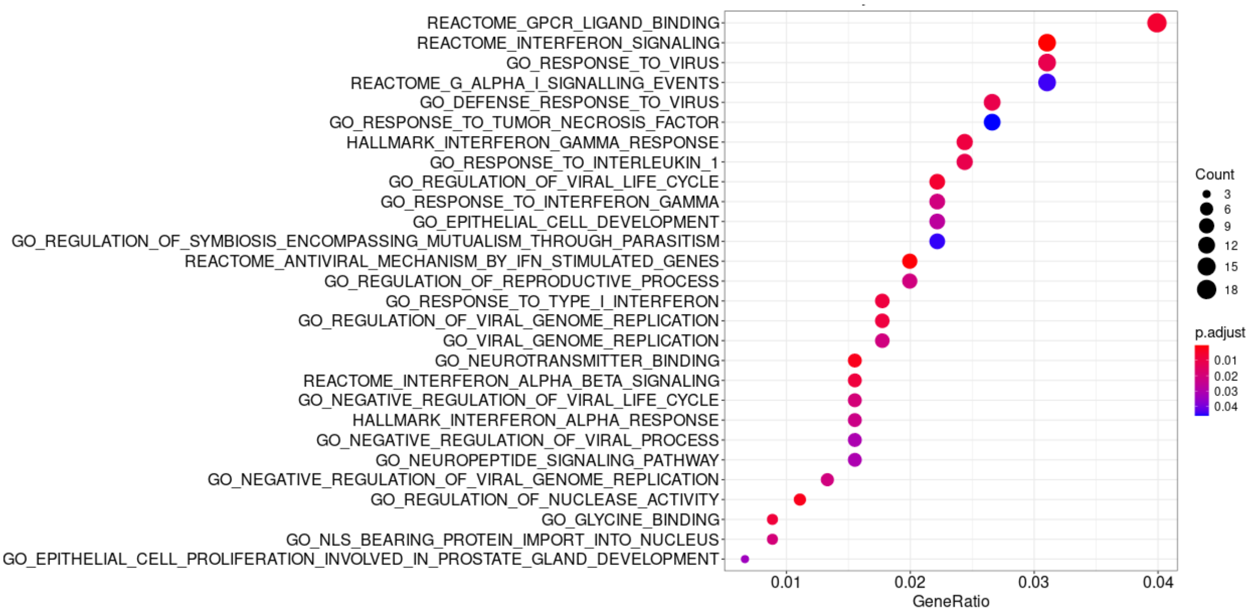


**B**

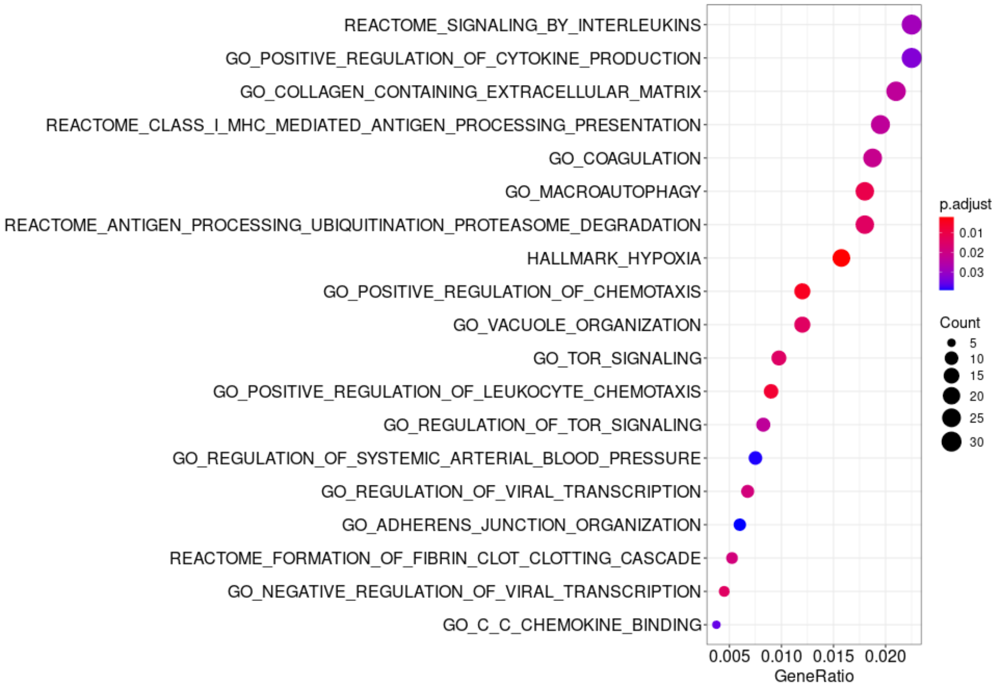

**Supplementary Figure 3.**

A.)

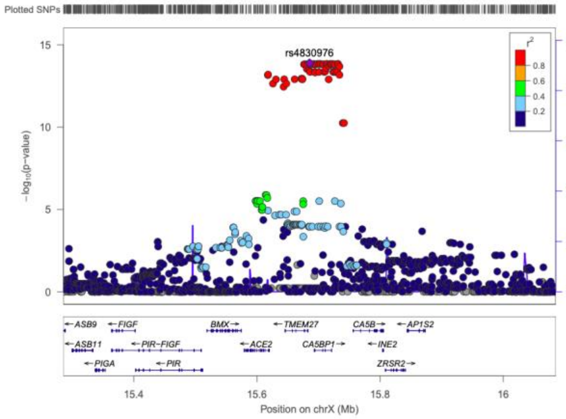


B.)

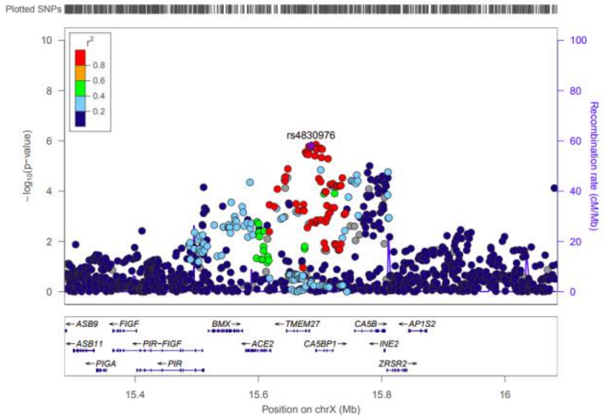


C.)

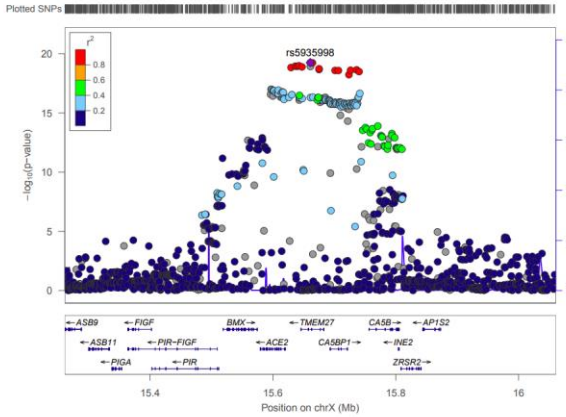


D.)

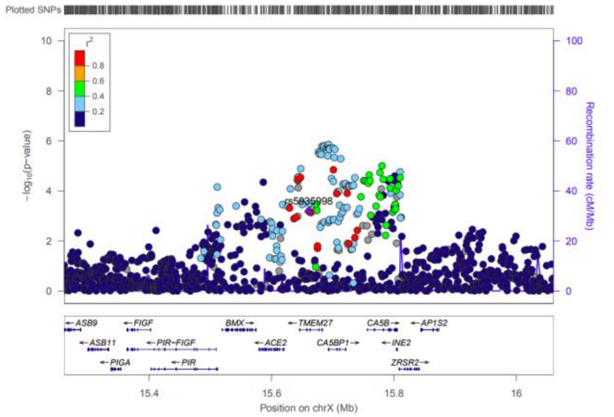


E.)

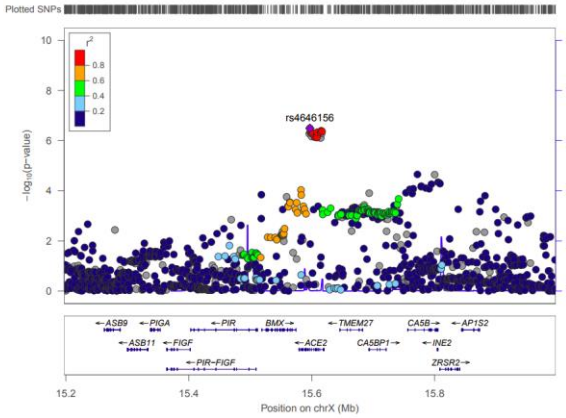


F.)

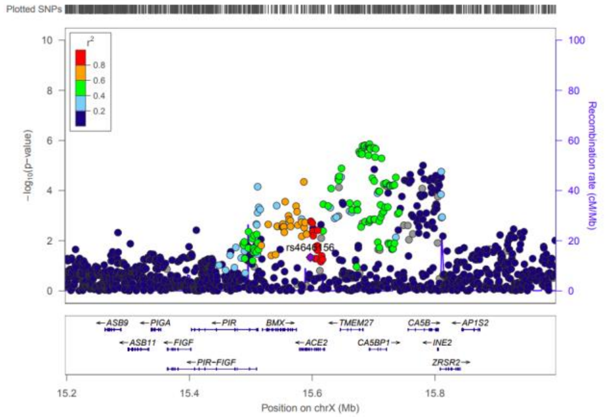
